# Genome-wide analysis in 756,646 individuals provides first genetic evidence that *ACE2* expression influences COVID-19 risk and yields genetic risk scores predictive of severe disease

**DOI:** 10.1101/2020.12.14.20248176

**Authors:** J. E. Horowitz, J. A. Kosmicki, A. Damask, D. Sharma, G. H. L. Roberts, A. E. Justice, N. Banerjee, M. V. Coignet, A. Yadav, J. B. Leader, A. Marcketta, D. S. Park, R. Lanche, E. Maxwell, S. C. Knight, X. Bai, H. Guturu, D. Sun, A. Baltzell, F. S. P. Kury, J. D. Backman, A. R. Girshick, C. O’Dushlaine, S. R. McCurdy, R. Partha, A. J. Mansfield, D. A. Turissini, A. H. Li, M. Zhang, J. Mbatchou, K. Watanabe, L. Gurski, S. E. McCarthy, H. M. Kang, L. Dobbyn, E. Stahl, A. Verma, G. Sirugo, Regeneron Genetics Center, M. D. Ritchie, M. Jones, S. Balasubramanian, K. Siminovitch, W. J. Salerno, A. R. Shuldiner, D. J. Rader, T. Mirshahi, A. E. Locke, J. Marchini, J. D. Overton, D. J. Carey, L. Habegger, M. N. Cantor, K. A. Rand, E. L. Hong, J. G. Reid, C. A. Ball, A. Baras, G. R. Abecasis, M. A. Ferreira

## Abstract

SARS-CoV-2 enters host cells by binding angiotensin-converting enzyme 2 (ACE2). Through a genome-wide association study, we show that a rare variant (MAF = 0.3%, odds ratio 0.60, *P*=4.5x10^-13^) that down-regulates *ACE2* expression reduces risk of COVID-19 disease, providing human genetics support for the hypothesis that ACE2 levels influence COVID-19 risk. Further, we show that common genetic variants define a risk score that predicts severe disease among COVID-19 cases.

## MAIN TEXT

Severe acute respiratory syndrome coronavirus 2 (SARS-CoV-2) causes coronavirus disease 2019 (COVID-19), which has lead to >3 million deaths worldwide since December 2019 ^1^. Reported risk factors for severe COVID-19, defined here as death or hospitalization combined with respiratory failure ^2^, include male sex, older age, race, obesity, kidney, cardiovascular and respiratory diseases ^3–5^. In this study, we used human genetics to identify genetic variants associated with severe COVID-19 and tested the utility of genetic risk scores to identify individuals at highest risk of severe disease.

We performed genome-wide association studies (GWAS) of COVID-19 outcomes across 52,630 individuals with COVID-19 and 704,016 individuals with no record of SARS-CoV-2 infection aggregated from four studies (Geisinger Health System [GHS], Penn Medicine BioBank [PMBB], UK Biobank [UKB] and AncestryDNA; **Supplementary Table 1**) and five continental ancestries. Of the COVID-19 cases, 6,911 (13.1%) were hospitalized and 2,184 (4.1%) had severe disease; hospitalized patients were more likely to be older, of non-European ancestry and to have pre-existing cardiovascular and lung disease (**Supplementary Table 2)**. Using these data, we defined two groups (risk and severity) of COVID-19 outcomes, ultimately resulting in five case-control comparisons related to risk of infection and two others related to disease severity among COVID-19 cases (**Table 1** and **Supplementary Table 3).** For each comparison, we performed ancestry-specific GWAS in each study and then combined results using a fixed-effects meta-analysis. Genomic inflation factors (λ_GC_) for the meta-analyses were <1.05, suggesting no substantial impact of population structure or unmodeled relatedness (**Supplementary Table 4).**

**Table 1.**
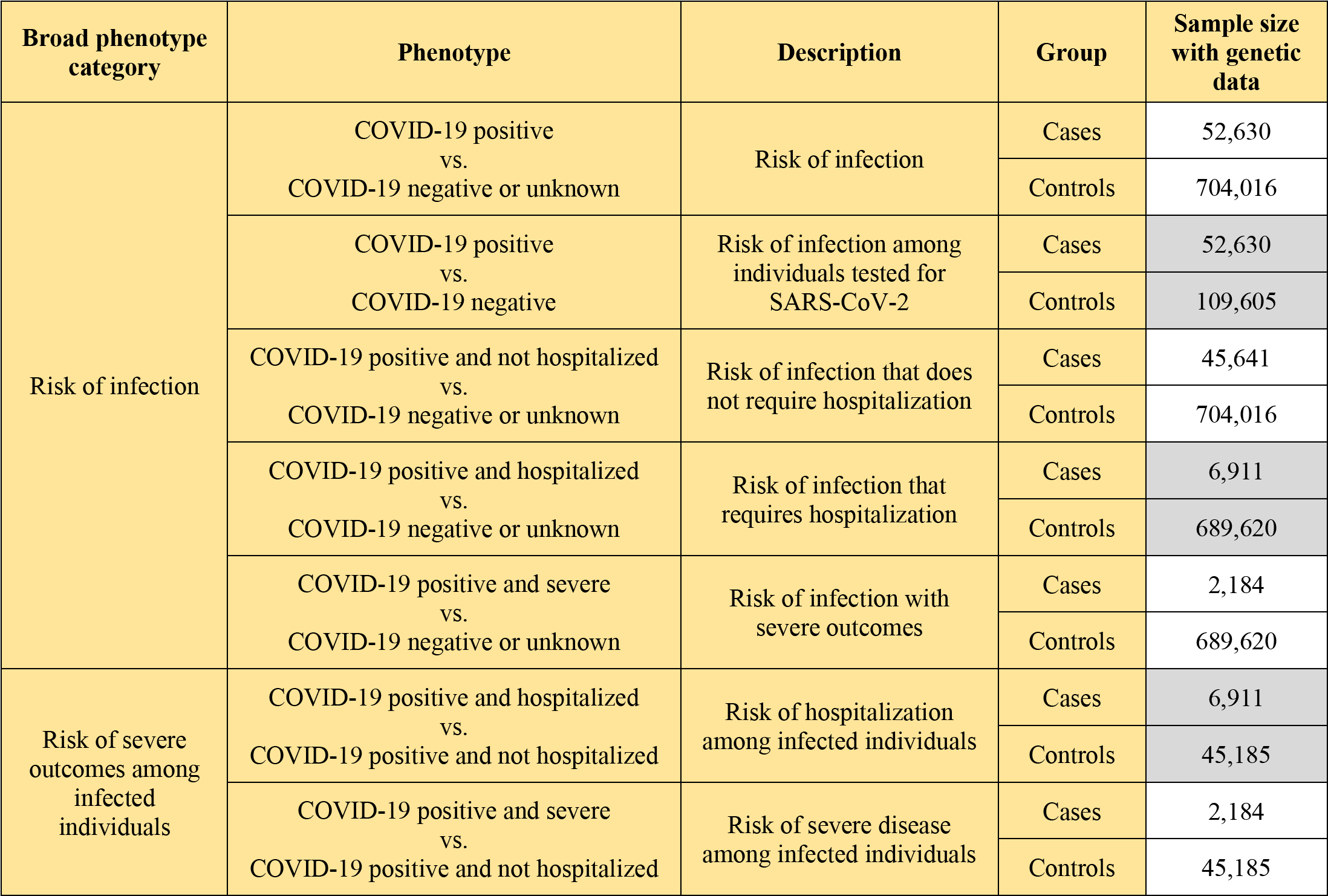
Seven COVID-19 phenotypes analyzed in this study.

Our analysis provides independent support for several risk variants reported in previous GWAS of COVID-19 (**Supplementary Table 5**), including those recently reported by the COVID-19 Host Genetics Initiative (HGI) ^6^, to which we contributed an earlier version of these data (**Supplementary Table 6**). Details for these replicated loci follow below, but first we looked for novel genetic associations that might have been missed by the HGI. Across the seven risk and severity phenotypes, considering both common (MAF>0.5%, up to 13 million) and rare (MAF<0.5%, up to 76 million) variants, we observed one previously unreported association at a conservative *P*<8x10^-11^ (Bonferroni correction for seven phenotypes x 89 million variants). This association was between lower risk of infection and rs190509934:C on the X-chromosome (MAF=0.3%, OR=0.60, 95% CI 0.52-0.69, *P* = 4.5x10^-13^; **Figure 1A and 1B**). This rare variant is located on the X chromosome, 60 base pairs upstream of the angiotensin-converting enzyme 2 gene (*ACE2*), the primary cell entry receptor for SARS-CoV-2 ^7^.

**Figure 1.**
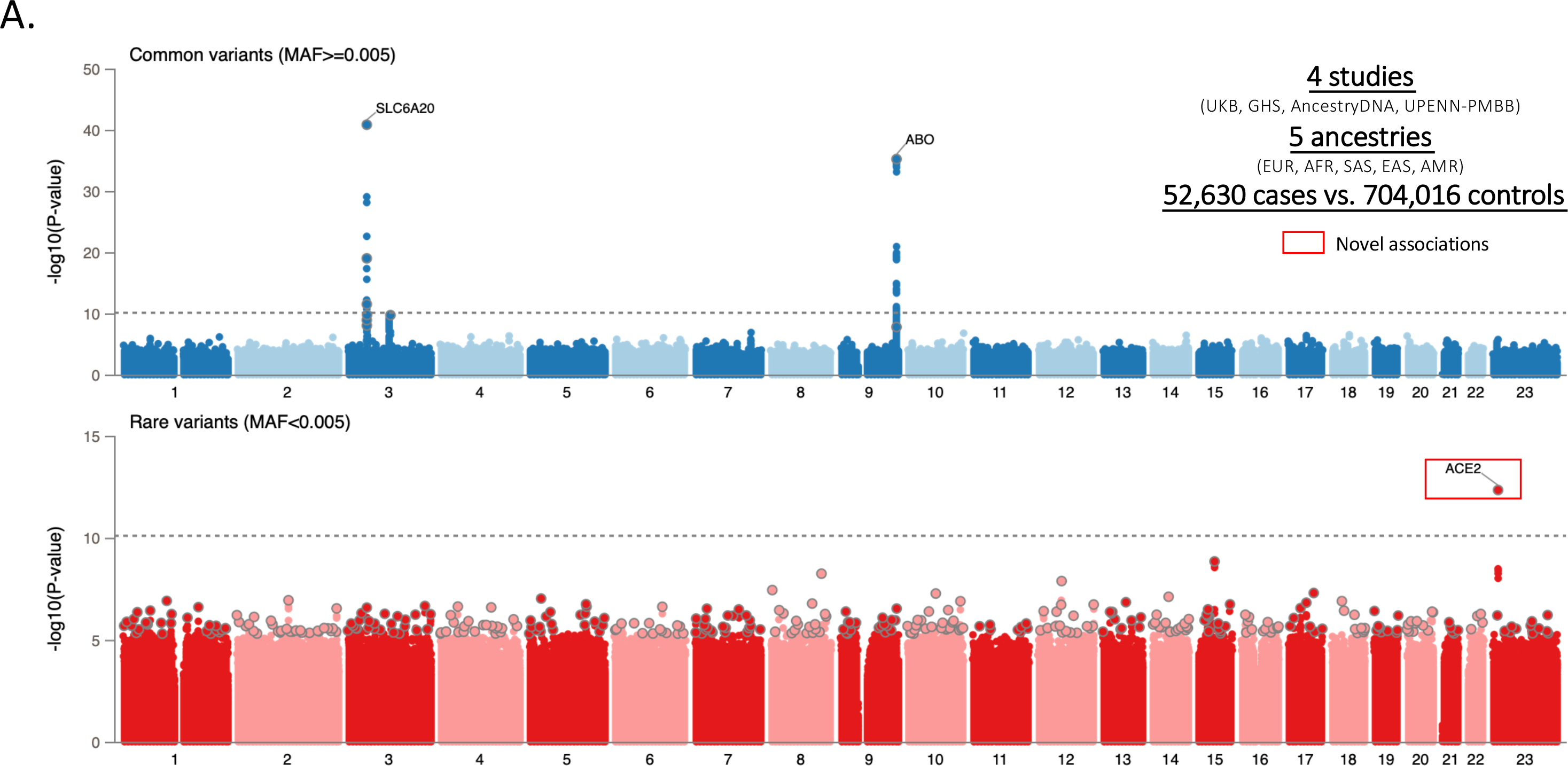
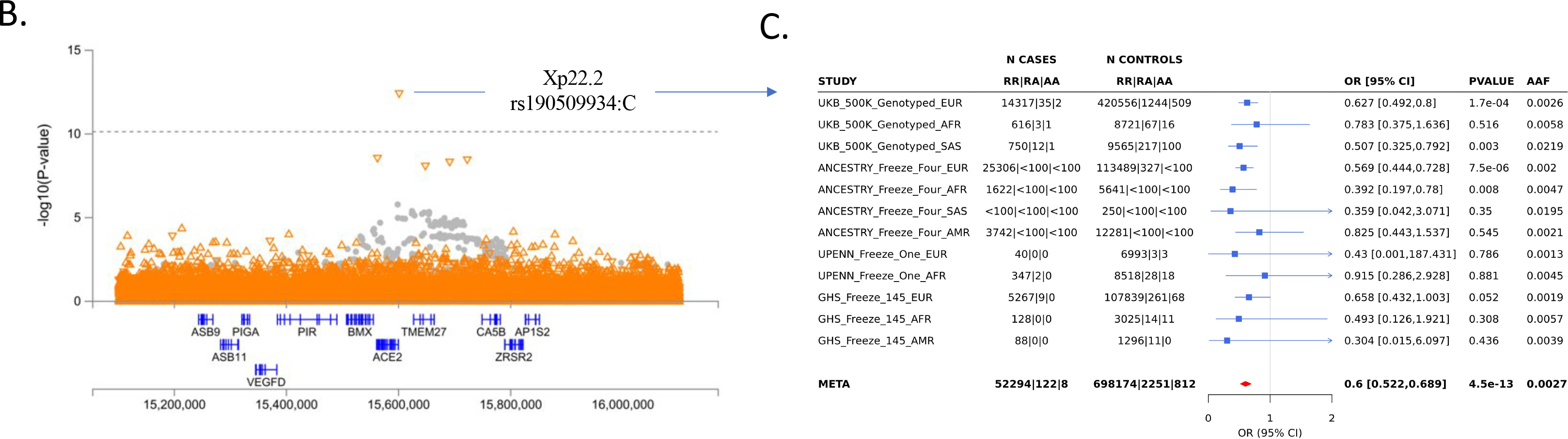
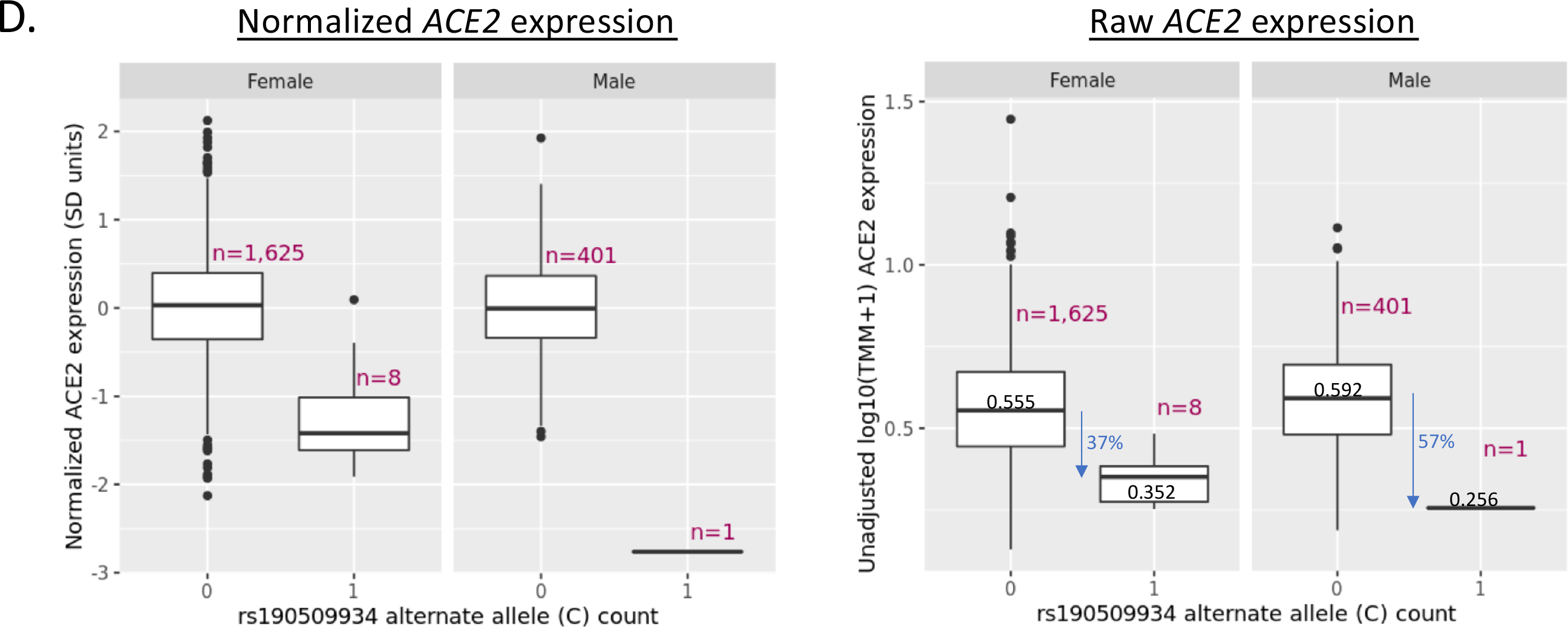
GWAS of 52,630 COVID-19 positive cases vs. 704,016 COVID-19 negative or unknown controls identifies a novel association with a rare variant near *ACE2* that lowers gene expression and protects against COVID-19. (**A**) Summary of association results for common (MAF>0.5%) and rare (MAF< 0.5%) variants. (**B**) Regional association plot centered around rs190509934 near ACE2. (**C**) Breakdown of association results across studies included in the meta-analysis of rs190509934. (**D**) Association between rs190509934:C and *ACE2* expression in liver measured in 2,035 individuals from the GHS study.

Given the potential significance of these findings, we studied the association between the *ACE2* variant rs190509934 and COVID-19 outcomes in greater detail. We found that the variant was well imputed (imputation info score >0.6 for all studies), and that there was no evidence for differences in effect size (heterogeneity test *P*>0.05) across studies (**Figure 1C**) or ancestries (**Supplementary Table 7**). However, a significantly stronger association (*P*=0.009) was observed in males (OR=0.49, *P*=7.0x10^-11^; explaining 0.085% of the variance in disease liability ^8^, *h*^2^) when compared to females (OR=0.72, *P*=5x10^-4^; *h*^2^=0.017%). There were no associations between rs190509934 and clinical risk factors for COVID-19 after correcting for multiple testing (**Supplementary Table 8**), suggesting that these were not likely confounders in the analysis. We then investigated the association between rs190509934 and severity among COVID-19 cases, and found that carriers of rs190509934:C had numerically (but not significantly) lower risk of worse disease outcomes when compared to non-carriers (**Supplementary Table 9**). These results demonstrate that rs190509934 near *ACE2* confers protection against SAR-CoV-2 infection and potentially also modulates disease severity among infected individuals; since the variant is rare, a definite account of its role on disease severity will require larger numbers of severe cases.

We speculated that the protective rare variant near *ACE2* (rs190509934:C) might regulate *ACE2* expression. This variant was not characterized by GTEx ^9^ or other gene expression studies we queried (**Supplementary Table 10**). Thus, to test its association with *ACE2* expression, we analyzed RNA-seq data from liver tissue available in a subset of 2,035 individuals from the GHS study, including eight heterozygous and one hemizygous carrier for rs190509934:C. After adjusting for potential confounders (e.g. body mass index, liver disease), we found that rs190509934:C reduced *ACE2* expression by 0.87 standard deviation units (95% CI -1.18 to -0.57, *P*=2.7x10^-8^; **Figure 1D**). When considering raw, pre-normalized *ACE2* expression levels, rs190509934:C was associated with a 39% reduction in expression relative to non-carriers. There was no association with the expression of 12 other nearby genes (within 500 kb) after accounting for multiple testing. These results are consistent with rs190509934:C lowering *ACE2* expression, which in turn confers protection from SARS-CoV-2 infection.

In addition to its role in viral infections, the normal physiologic role of ACE2 involves its hydrolysis and clearance of angiotensin II, a vasoconstrictive peptide that can lead to higher vascular tone or blood pressure ^10^. Therefore, we investigated if rs190509934:C was associated with higher systolic blood pressure in the UKB study, but found no significant association (beta=0.009 SD-units, *P*=0.56; **Supplementary Table 11**). There was a trend for higher blood pressure among carriers of ultra-rare coding variants in *ACE2* that are predicted to be full loss-of-function (beta=0.219, *P*=0.086; **Supplementary Table 11**), assayed through exome-sequencing as described previously ^11^. These results need to be confirmed in larger datasets, but suggest that ACE2 loss-of-function may modestly increase blood pressure. This should be considered if ACE2 blockade is to be developed for COVID-19 treatment, although pharmacologic inhibition of ACE2 in such a setting would be expected to be short term and elevations in blood pressure could be managed with anti-hypertensives. Of note, *ACE2* expression in the airways was reported to be higher in smokers and patients with COPD ^12^ and to increase with age ^13^. Collectively, these observations and our genetic findings are consistent with the hypothesis that ACE2 levels play a key role in determining COVID-19 risk.

As noted, our GWAS also identified associations at several loci reported in previous GWAS of COVID-19 outcomes. To explore previously reported signals in detail, we first attempted to replicate eight independent associations (*r*^2^<0.05) with disease risk (**Supplementary Table 5)** reported in three recent GWAS that included >1,000 cases (**Supplementary Table 6**). After accounting for multiple testing, six variants had a significant (*P*<0.0012) and directionally consistent association in at least one of our five disease risk analyses (**Supplementary Table 12**), specifically those located in/near *LZTFL1*, *SLC6A20,* MHC, *ABO, DPP9* and *IFNAR2.* There was no evidence for heterogeneity in effect sizes across studies (**Supplementary Table 12**) or ancestries (**Supplementary Table 13**). We also explored the possibility that the association between these six variants and COVID-19 risk could have been confounded by disease status for relevant comorbidities. We found that only two of the six variants were modestly associated with a clinical risk factor (**Supplementary Table 8**), and we conclude that it is unlikely that the association between the six variants and risk of COVID-19 is explained by these underlying comorbidities.

To evaluate whether genetics could be used to predict severe disease, we first investigated which replicated variants were associated with severity amongst COVID-19 cases. Among the six replicated variants (in/near *LZTFL1*, *SLC6A20*, MHC, *ABO*, *DPP9* and *IFNAR2*), four were significantly (*P*<0.05) associated with worse outcomes among infected individuals (in/near *LZTFL1*, MHC, *DPP9* and *IFNAR2*), while those in *ABO* and near *SLC6A20* did not associate with COVID-19 severity (**Supplementary Figure 1B** and **Supplementary Table 14**). Collectively, these results highlight four variants associated with both COVID-19 risk and worse disease outcomes, including respiratory failure and death. These variants may be used to identify individuals at risk of severe COVID-19 and to guide the search for genes involved in the pathophysiology of COVID-19.

We next evaluated whether variants identified by the COVID-19 HGI, a large worldwide effort to identify genetic risk factors for COVID-19, could augment this set of four disease severity variants. The latest HGI analyses^6^ include data from 49,562 SARS-CoV-2 infected individuals and use >1.7 million individuals with no record of infection as controls (**Supplementary Table 15**). To identify additional variants associated with severity, we started with variants associated with the phenotype “reported infection” (infected vs. no record of infection) which, despite the sample overlap between the HGI and our analyses, is statistically independent from severity among infected individuals – since infection status (positive vs. negative or unknown) is uncorrelated with hospitalization status once infected (hospitalized vs. not hospitalized). We found that two were nominally associated with risks of hospitalization or severe disease among cases (rs11919389 near *RPL24* and rs1886814 near *FOXP4*; **Supplementary Table 15**), suggesting that these loci also modulate disease severity after infection with SARS-CoV-2.

Collectively, our association analyses highlighted six common variants identified in previous GWAS or by the HGI - in/near *LZTFL1*, MHC, *DPP9*, *IFNAR2*, *RPL24* and *FOXP4* - that are associated with risk of COVID-19 as well as disease severity among cases. To help identify genes that might underlie the observed associations, we searched for functional protein-coding variants (missense or predicted loss-of-function) in high LD (*r*^2^>0.80) with each variant. We found eight functional variants in five genes (**Supplementary Table 16**): *IFNAR2*, a cytokine receptor component in the anti-viral type 1 interferon pathway, which is activated by SARS-CoV-2 and is dysregulated in severe COVID-19 cases ^14–16^; *CCHCR1*, a P-body protein associated with cytoskeletal remodeling and mRNA turnover ^17, 18^; *TCF19*, a transcription factor associated with hepatitis B ^19^; and *C6orf15* and *PSORS1C1*, two functionally uncharacterized genes in the MHC. These data indicate that the risk variants identified may have functional effects on these five genes.

Next, we asked if any of the six sentinel risk variants co-localized (*i.e.* were in high LD, *r*^2^>0.80) with published sentinel expression quantitative trait loci (eQTL) across 52 studies (**Supplementary Table 10**), specifically focusing on 114 genes in *cis* (± 500 kb). We found co-localization with sentinel eQTL for eight genes (**Supplementary Table 17)**: *SLC6A20* (eQTL from lung), a proline transporter that binds the host SARS-CoV-2 receptor, ACE2 ^20^; *NXPE3* (esophagus), a gene of uknown function; *SENP7* (blood), a SUMO-specific protease that promotes interferon signaling and that in mice is essential for innate defense against herpes simplex virus 1 infection^21^; *IFNAR2* and *TCF19* (multiple tissues), both discussed above*; LST1* (blood), an immune modulatory protein that inhibits lymphocyte proliferation ^22^ and is upregulated in response to bacterial ligands ^23^; *HLA-C* (adipose), a natural killer cell ligand, associated with viral infection ^24^ and autoimmunity ^25^; and *IL10RB* (multiple tissues), a pleotropic cytokine receptor associated with persistent hepatitis B and autoimmunity ^26, 27^.

Collectively, analysis of missense variation and eQTL catalogs suggests 12 potential effector genes in COVID-19 loci (*ACE2*, *C6orf15*, *CCHCR1*, *HLA*-C, *IFNAR2*, *IL10RB*, *LST1*, *NXPE3*, *PSORS1C1, SENP7*, *SLC6A20* and *TCL19*), though functional studies are required to confirm these predictions.

Next, we proceeded to evaluate if common genetic variants can help identify individuals at high risk of severe disease. Specifically, we focused on the six variants (in/near *LZTFL1*, MHC, *DPP9*, *IFNAR2, RPL24* and *FOXP4*) that associated with both risk of COVID-19 and disease severity among infected individuals. Using these variants, we created a weighted genetic risk score (GRS) for individuals with COVID-19 and then compared the risk of hospitalization and severe disease between those with a high GRS and all other cases, after adjusting for established risk factors. The weights used for each variant corresponded to the effect size (log of the odds ratio) reported in previous GWAS. When considering COVID-19 cases of European ancestry (N=44,958), we found that having a high GRS (top 10%) was associated with a 1.38-fold increased risk of hospitalization (95% CI 1.26 to 1.53, *P*=6x10^-11^; **Figure 2A**) and 1.58-fold increased risk of severe disease (95% CI 1.36 to 1.82, *P*=7x10^-10^; **Figure 2B**). In other ancestries, a high GRS also appeared to predict risk of hospitalization – including among individuals of African ancestry (N=2,598, 1.70-fold risk for high GRS, 95% CI 1.03 to 2.81, *P*=0.038), Admixed American ancestry (N=3,752, 1.56-fold risk, 95% CI 1.00 to 2.43, *P* = 0.05) and South Asian ancestry (N=760, 1.42-fold risk, 95% CI 0.72 to 2.82, *P* = 0.32, **Supplementary Table 18**). A similar pattern was observed for risk of severe disease, though sample sizes were considerably smaller (**Supplementary Tables 19**).

**Figure 2.**
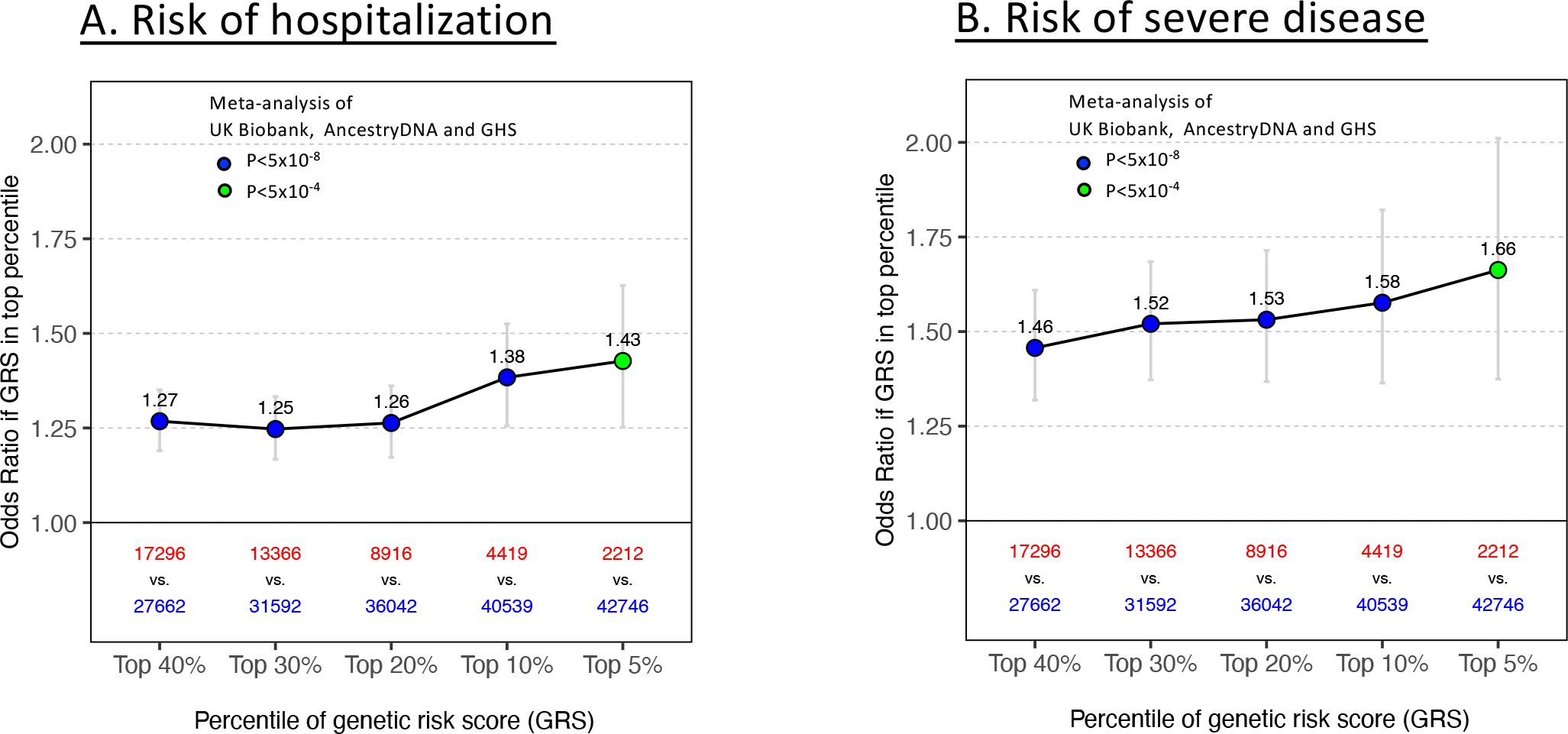
Association between a 6-SNP genetic risk score (GRS) and risk of hospitalization (A) and severe disease (B) among COVID-19 cases of European ancestry. (**A**) Association between high genetic risk and hospitalization. Risk of hospitalization among cases is shown for individuals in the top GRS percentile, agnostic to the number of clinical risk factors present. The association was tested in three studies separately (AncestryDNA, UKB and GHS studies) using logistic regression, with established risk factors for COVID-19 included as covariates (see Methods for details). Results were then meta-analyzed across studies, for a combined sample size of 44,958 COVID-19 cases, including 6,138 hospitalized. N in red: number of COVID-19 cases in the top GRS percentile. Error bars represent 95% confidence intervals. (**B**) Association between high genetic risk and severe disease. The association was tested as described above in three studies separately (AncestryDNA, UKB and GHS studies). Results were then meta-analyzed across studies, for a combined sample size of 44,958 COVID-19 cases, including 1,940 with severe disease. N in red: number of COVID-19 cases in the top GRS percentile. N in grey: number of COVID-19 cases in the rest of population.

We then compared the effect of the GRS between individuals with and without established risk factors for severe COVID-19. In Europeans of both the AncestryDNA and UK Biobank studies, we found that a high GRS (top 10%) was associated with risk of severe disease both among individuals with and without established clinical risk factors for severe COVID-19 (**Figure 3**). In the meta-analysis of the two studies, a high GRS was associated with a 1.65-fold (95% CI 1.39-1.96, *P*=1x10^-8^) and 1.75-fold (95% CI 1.28-2.40, *P*=4x10^-4^) higher risk of severe disease, respectively among individuals with and without established risk factors (**Supplementary Table 20**). There was no evidence for heterogeneity of GRS effect with clinical risk factor status (*P*=0.30). Similar results were observed for (i) risk of hospitalization (**Supplementary Figure 2** and **Supplementary Table 20**); (ii) when including in the GRS all 12 variants reported to associate with risk of COVID-19 in previous GWAS (eight variants) and by the HGI (four novel variants associated with reported infection; **Supplementary Figure 3**); and (iii) in individuals of Admixed American ancestry (**Supplementary Figure 4**; stratitifed analysis not performed in other ancestries due to small sample size). Lastly, we also found that expanding the GRS to include a larger set of variants did not improve the observed associations (**Supplementary Figure 5**). Overall, these results demonstrate that a GRS calculated using variants associated with disease risk and severity can potentially be used to identify COVID-19 cases at high risk of developing poor disease outcomes. We note that preeminent factor for severe COVID-19 outcomes remains age (as illustrated in **Supplementary Figure 6**), but that common genetic variants appear to provide complementary information that may be used to stratify risk among older individuals.

**Figure 3.**
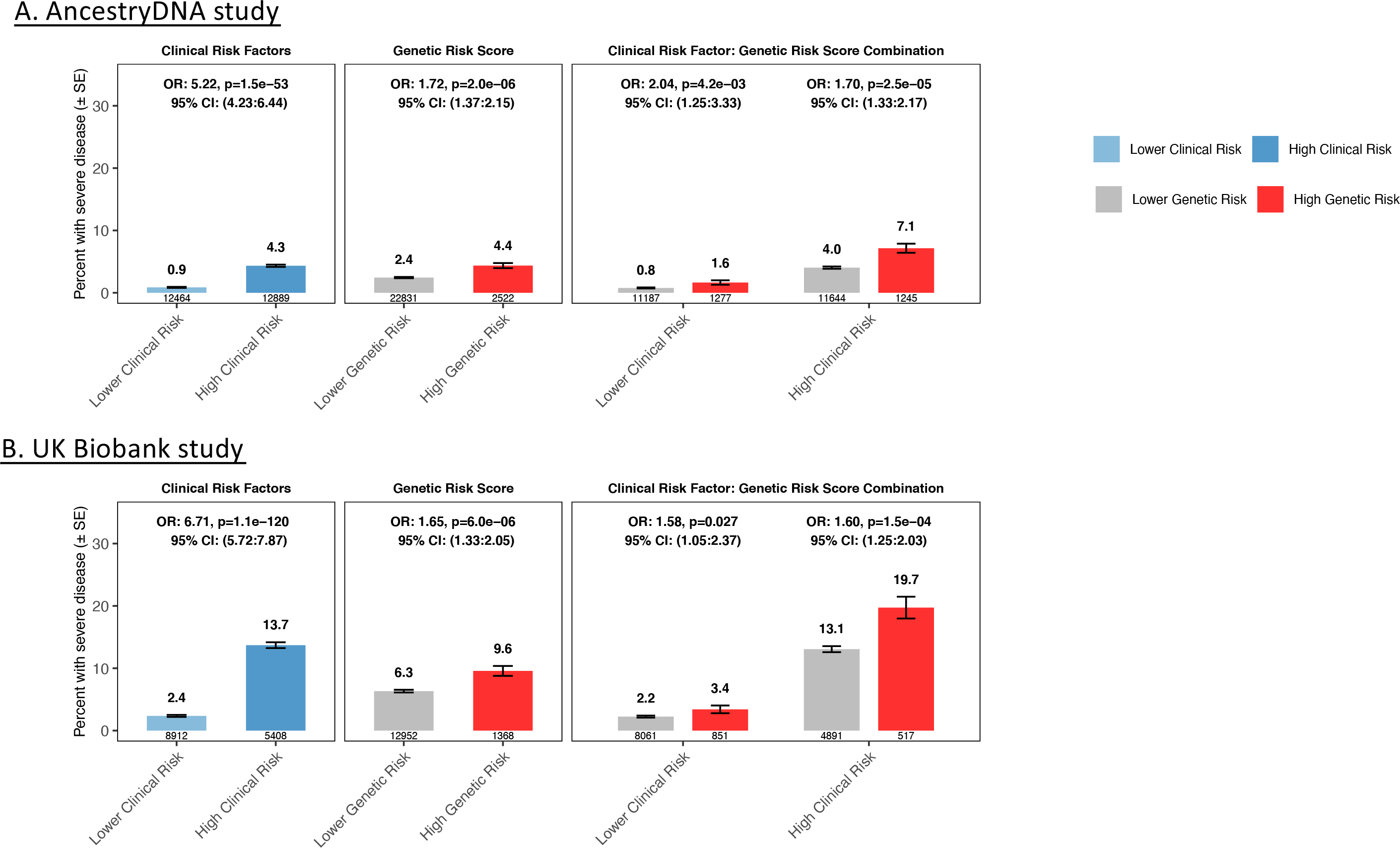
Association between a 6-SNP genetic risk score (GRS) and risk of severe disease among COVID-19 cases of European ancestry after stratifying by the presence of clinical risk factors. (**A**) Rate of severe disease in the AncestryDNA study (25,353 COVID-19 cases, including 667 with severe disease). (**B**) Rate of severe disease in the UK Biobank study (14,320 COVID-19 cases, including 951 with severe disease). High genetic risk (red bars): top 10% of the GRS. Low genetic risk (grey bars): bottom 90% of the GRS (*i.e*. all other COVID-19 cases). Error bars (black) represent 95% confidence intervals.

The following caveats should be considered when interpreting results from this study. First, our study had greater power to identify associations with disease risk than with severity outcomes, given the relatively small sample size for the latter. Second, there was phenotypic heterogeneity among COVID-19 cases and controls and associated risk factors across our studies. One likely reason for this is that survey respondents from the AncestryDNA study were enriched for healthier individuals and milder COVID-19 cases, when compared to participants of the UKB, GHS and PMBB studies, who were ascertained in clinical settings and so were enriched for hospitalized and severe COVID-19 cases. Other sources of heterogeneity may include regional and temporal availability of COVID-19 testing and the inability to control for viral exposure among controls. While our meta-analysis collectively spans a broad phenotypic spectrum, these individual differences may account for variability in results across reported studies. Third, we used expression levels measured in liver to assess the impact of the ACE2 risk variant on gene expression. Liver is not the most relevant tissue to assess *ACE2* expression, but we note that cis eQTLs are often shared across tissues ^9, 28^. Fourth, the association between GRS and risk of severe disease was strongest in European individuals of the AncestryDNA (OR=1.72, *P*=2x10^-6^) and UKB (OR=1.65, *P*=6x10^-6^) studies when compared to the smaller GHS study (OR=1.03, *P*=0.877). The lower effect size in the latter may be due to differences in ascertainment of COVID-19 positive cases, as discussed above, or stochastic, given the smaller sample size. We also noted that the impact of the GRS on risk of hospitalization was attenuated in comparison to severe disease, which may be a reflection of the weighting schema for the variants comprising the score; the four largest GRS weights were derived from an analysis of critically ill individuals.

In summary, we confirmed six common variant associations with risk of infection and further show that four of these variants modulate disease severity among cases. We also identified one novel association with disease risk which provides human genetic support for the hypothesis that ACE2 expression plays a key role SARS-CoV-2 infection and may constitute an attractive therapeutic target for prevention COVID-19 disease and its sequelae. Lastly, we demonstrate that a genetic risk score based on common variants validated in this study can be used to identify individuals at high risk of poor disease outcomes.

## ONLINE METHODS

### Participating Studies

#### AncestryDNA COVID-19 Research Study

AncestryDNA customers over age 18, living in the United States, and who had consented to research, were invited to complete a survey assessing COVID-19 outcomes and other demographic information. These included SARS-CoV-2 swab and antibody test results, COVID-19 symptoms and severity, brief medical history, household and occupational exposure to SARS-CoV-2, and blood type. A total of 163,650 AncestryDNA survey respondents were selected for inclusion in this study ^29^. Respondents selected for this study included all individuals with a positive COVID-19 test together with age and sex matched controls. DNA samples were genotyped as described previously^29^. Genotype data for variants not included in the array were then inferred using imputation to the Haplotype Reference Consortium (HRC) reference panel. Briefly, samples were imputed to HRC version 1.1, which consists of 27,165 total individuals and 36 million variants. The HRC reference panel does not include indels; consequently, indels are not present in the imputed data. We determined best-guess haplotypes with Eagle version 2.4.1 and performed imputation with Minimac4 version 1.0.1. We used 1,117,080 unique variants as input and 8,049,082 imputed variants were retained in the final data set. Variants with a Minimac4 R^2^<0.30 were filtered from the analysis.

#### Geisinger Health System (GHS)

The GHS MyCode Community Health Initiative is a health system-based cohort from central and eastern Pennsylvania (USA) with ongoing recruitment since 2006^30^. A subset of 144,182 MyCode participants sequenced as part of the GHS-Regeneron Genetics Center DiscovEHR partnership were included in this study. Information on COVID-19 outcomes were obtained through GHS’s COVID-19 registry. Patients were identified as eligible for the registry based on relevant lab results and ICD-10 diagnosis codes; patient charts were then reviewed to confirm COVID-19 diagnoses. The registry contains data on outcomes, comorbidities, medications, supplemental oxygen use, and ICU admissions. DNA from participants was genotyped on either the Illumina OmniExpress Exome (OMNI) or Global Screening Array (GSA) and imputed to the TOPMed reference panel (stratified by array) using the TOPMed Imputation Server. Prior to imputation, we retained variants that had a MAF >= 0.1%, missingness < 1% and HWE p-value > 10^-15^. Following imputation, data from the OMNI and GSA datasets were merged for subsequent association analyses, which included an OMNI/GSA batch covariate, in addition to other covariates described below.

#### Penn Medicine BioBank (PMBB) study

PMBB contains ∼70,000 study participants, all recruited through the University of Pennsylvania Health System (UPHS). Participants donate blood or tissue and allow access to EHR information ^31^. The PMBB participants with COVID-19 infection were identified through the UPHS COVID-19 registry, which consists of qPCR results of all patients tested for SARS-CoV-2 infection within the health system. We then used electronic health records to classify COVID-19 patients into hospitalized and severe (ventilation or death) categories. DNA genotyping was performed with the Illumina Global Screening Array, and imputation performed using the TOPMed reference panel as described for GHS above.

#### UK Biobank (UKB) study

We studied the host genetics of SARS-CoV-2 infection in participants of the UK Biobank study, which took place between 2006 and 2010 and includes approximately 500,000 adults aged 40-69 at recruitment. In collaboration with UK health authorities, the UK Biobank has made available regular updates on COVID-19 status for all participants, including results from four main data types: qPCR test for SARS-CoV-2, anonymized electronic health records, primary care and death registry data. We report results based on phenotype data downloaded on the 4th January 2021 and excluded from the analysis 28,547 individuals with a death registry event prior to 2020. DNA samples were genotyped as described previously ^32^ using the Applied Biosystems UK BiLEVE Axiom Array (N=49,950) or the closely related Applied Biosystems UK Biobank Axiom Array (N=438,427). Genotype data for variants not included in the arrays were inferred using the TOPMed reference panel, as described above.

### COVID-19 phenotypes used for genetic association analyses

We grouped participants from each study into three broad COVID-19 disease categories (**Supplementary Table 1**): (i) positive – those with a positive qPCR or serology test for SARS-CoV-2, or with a COVID-19-related ICD10 code (U07), hospitalization or death; (ii) negative – those with only negative qPCR or serology test results for SARS-CoV-2 and with no COVID-19-related ICD10 code (U07), hospitalization or death; and (iii) unknown – those with no qPCR or serology test results and no COVID-19-related ICD10 code (U07), hospitalization or death. We then used these broad COVID-19 disease categories, in addition to hospitalization and disease severity information, to create seven COVID-19-related phenotypes for genetic association analyses, as detailed in **Supplementary Table 3**.

SARS-CoV-2 infection status (positive, negative or unknown) was determined based on a qPCR test for SARS-CoV-2 in the UKB, GHS and PMBB studies; self-reported results for qPCR or serology test for SARS-CoV-2 in the AncestryDNA study.

Hospitalization status (positive, negative or unknown) was determined based on COVID-19-related ICD10 codes U071, U072, U073 in variable ‘diag_icd10’ (table ‘hesin_diag’) in the UKB study; self-reported hospitalization due to COVID-19 in the AncestryDNA study; medical records in the GHS and PMBB studies.

Disease severity status (severe [ventilation or death] or not severe) was determined in the UKB study based on (i) respiratory support ICD10 code Z998 in variable ‘diag_icd10’ (table ‘hesin_diag’); (ii) the following respiratory support ICD10 codes in variable ‘oper4’ (table ‘hesin_oper’): E85, E851, E852, E853, E854,E855, E856, E858, E859, E87, E871, E872, E873, E874, E878, E879, E89, X56, X561, X562, X563, X568, X569, X58, X581, X588, X589; or (3) COVID-19-related ICD10 codes U071, U072, U073 in cause of death (variable ‘cause_icd10’ in table ‘death_cause’). In the AncestryDNA study, disease severity was determined based on self-reported ventilation or need for supplementary oxygen due to COVID-19. In the GHS and PMBB study, it was determined based on ventilator or high-flow oxygen use.

For association analysis in the AncestryDNA study, we excluded from the COVID-19 unknown group individuals who had (i) a first-degree relative who was COVID-19 positive; or (ii) flu-like symptoms.

### Genetic association analyses

Association analyses in each study were performed using the genome-wide Firth logistic regression test implemented in REGENIE ^33^. In this implementation, Firth’s approach is applied when the p-value from standard logistic regression score test is below 0.05. We included in step 1 of REGENIE (*i.e.* prediction of individual trait values based on the genetic data) directly genotyped variants with a minor allele frequency (MAF) >1%, <10% missingness, Hardy-Weinberg equilibrium test *P*-value>10^-15^ and linkage-disequilibrium (LD) pruning (1000 variant windows, 100 variant sliding windows and *r*^2^<0.9). The association model used in step 2 of REGENIE included as covariates age, age^2^, sex, age-by-sex, and the first 10 ancestry-informative principal components (PCs) derived from the analysis of a stricter set of LD-pruned (50 variant windows, 5 variant sliding windows and *r*^2^<0.5) common variants from the array (imputed for the GHS study) data.

Within each study, association analyses were performed separately for five different continental ancestries defined based on the array data: African (AFR), Admixed American (AMR), European (EUR) and South Asian (SAS). We determined continental ancestries by projecting each sample onto reference principle components calculated from the HapMap3 reference panel. Briefly, we merged our samples with HapMap3 samples and kept only SNPs in common between the two datasets. We further excluded SNPs with MAF<10%, genotype missingness >5% or Hardy-Weinberg Equilibrium test p-value < 10^-5^. We calculated PCs for the HapMap3 samples and projected each of our samples onto those PCs. To assign a continental ancestry group to each non-HapMap3 sample, we trained a kernel density estimator (KDE) using the HapMap3 PCs and used the KDEs to calculate the likelihood of a given sample belonging to each of the five continental ancestry groups. When the likelihood for a given ancestry group was >0.3, the sample was assigned to that ancestry group. When two ancestry groups had a likelihood >0.3, we arbitrarily assigned AFR over EUR, AMR over EUR, AMR over EAS, SAS over EUR, and AMR over AFR. Samples were excluded from analysis if no ancestry likelihoods were >0.3, or if more than three ancestry likelihoods were > 0.3.

Results were subsequently meta-analyzed across studies and ancestries using an inverse variance-weighed fixed-effects meta-analysis.

### Identification of putative targets of GWAS variants based on colocalization with eQTL

We identified as a likely target of a sentinel GWAS variant any gene for which a sentinel expression quantitative trait locus (eQTL) co-localized (*i.e.* had LD *r*^2^ > 0.80) with the sentinel GWAS variant. That is, we only considered genes for which there was strong LD between a sentinel GWAS variant and a sentinel eQTL, which reduces the chance of spurious colocalization. Sentinel eQTL were defined across 174 published datasets (**Supplementary Table 10**), as described previously ^34^. We did not use statistical approaches developed to distinguish colocalization from shared genetic effects because these have very limited resolution at high LD levels (*r*^2^ > 0.80) ^35^.

### Gene expression analysis in participants of the GHS study

For a subset of individuals from the GHS study (n=2,035, ascertained through the Geisinger Bariatric Surgery Clinic), RNA was extracted from liver biopsies conducted during bariatric surgery to evaluate liver disease. Individuals had class 3 obesity (BMI>40kg/m^2^) or class 2 obesity (BMI 35-39 kg/m^2^) with an obesity-related co-morbidity (e.g. type-2 diabetes, hypertension, sleep apnea, non-alcoholic fatty liver disease). RNA libraries were prepared using polyA-extraction and then sequenced with 75bp paired-end reads with two 10 bp index reads on the Illumina NovaSeq 6000 on S4 flow cells. RNA-seq data were then analyzed using the GTEx v8 workflow^36^, using STAR ^37^ and RNASeqQC ^38^, except that GENCODE v32 was used in lieu of v26. Briefly: (i) raw expression counts were normalized with TMM (Trimmed Mean of M-values) as implemented in edgeR ^38^; (ii) a rank-based inverse normal transformation was applied to the normalized expression values; (iii) principal components (PCs) analysis was performed on data from 25,078 genes with TPM >0.1 in >20% samples, to identify latent factors accounting for variation in gene expression; (iv) gene expression levels were adjusted for the top 100 PCs to improve power to identify cis-regulatory effects. The association between adjusted *ACE2* expression and the imputed genotypes of rs190509934 was then tested using REGENIE, with the following variables included as covariates: age, age^2^, four ancestry-informative principal components, steatosis status, fibrosis status, diabetes status, and body mass index at the time of bariatric surgery.

### Genetic risk score (GRS) analysis of COVID-19 hospitalization and severity

First, in each study (AncestryDNA, GHS, UKB and PMBB), we created a GRS for each COVID-19 positive individual based on variants that were reported to associate with risk of COVID-19 in previous GWAS and that we (i) independently replicated (except variants identified by the HGI); and (ii) found to be associated with COVID-19 severity outcomes. We used as weights the effect (beta) reported in previous GWAS (**Supplementary Table 5**). Second, we ranked COVID-19 individuals based on the GRS and created a new binary GRS predictor by assigning each individual to a high (top 5%) or low (rest of the population) percentile group. Third, for studies with >100 hospitalized cases, we used logistic regression to test the association between the binary GRS predictor and risk of hospitalization (hospitalized cases vs. all other cases), including as covariates age, sex, age-by-sex interaction, and ten ancestry-informative PCs. In addition to age and sex, we included as additional covariates established clinical risk factors for COVID-19 that are outlined in the Emergency Use Authorisation treatment guidelines for casirivimab and imdevimb: BMI, chronic kidney disease, diabetes, immunosuppressive disease, chronic obstructive pulmonary disease or other chronic respiratory disease, cardiovascular disease and hypertension. We repeated the association analysis (i) using different percentile cut-offs for the GRS (5%, 10%, 20%, 30% and 40%); and (ii) to test the association with disease severity (severe cases vs. all other cases). We then stratified COVID-19 cases by clinical risk (high versus lower) and evaluated the association between the top 10% by GRS (i.e. high genetic risk) and risk of hospitalization or severe disease. The stratified analyses were performed with logistic regression, with sex and ancestry-informative PCs included as covariates. High clinical risk was defined as any one of the following: (i) age≥65; (ii) BMI≥35; (iii) chronic kidney disease, diabetes or immunosuppressive disease; (iv) age ≥55 and presence of chronic obstructive pulmonary disease/other chronic respiratory disease, cardiovascular disease, or hypertension.

## Code availability

Upload Agent (v1.5.30) can be found at https://wiki.dnanexus.com/Downloads#Upload-Agent.bcl2fastq software (v2.19.0) can be found at https://support.illumina.com/sequencing/sequencing_software/bcl2fastq-conversion-software.html. BWA software (v0.7.17) for read alignment can be found at http://bio-bwa.sourceforge.net. Picard software (v1.141) can be found at https://broadinstitute.github.io/picard/. Samtools (v1.7) can be found at http://www.htslib.org. WeCall (v1.1.2) can be found at https://github.com/Genomicsplc/wecall. FastQC (v0.11.8) can be found at http://www.bioinformatics/babraham.ac.uk/projects/fastqc/. Bcftools, bgzip, and tabix (v1.7) can be found at http://www.htslib.org, bgzip/tabix v1.7. pigz (v2.3.4) can be found at https://zlib.net/pigz/. Eagle (v2.4.1) can be found at https://github.com/poruloh/Eagle. Minimac4 (v1.01) can be found at https://github.com/statgen/Minimac4. GLnexus (v0.4.0) can be found at https://github.com/dnanexus-rnd/GLnexus. PLINK (v1.90b6.21) can be found at https://www.cog-genomics.org/plink2/. PRIMUS can be found at https://primus.gs.washington.edu/primusweb/. REGENIE (v2.0.1) can be found at https://github.com/statgen/METAL.

## Data availability

All genotype-phenotype association results reported in this study are available for browsing using the RGC’s COVID-19 Results Browser (https://rgc-covid19.regeneron.com). Data access and use is limited to research purposes in accordance with the Terms of Use (https://rgc-covid19.regeneron.com/terms-of-use).

## Data Availability

All genotype-phenotype association results reported in this study are available for browsing using the RGC COVID-19 Results Browser (https://rgc-covid19.regeneron.com). Data access and use is limited to research purposes in accordance with the Terms of Use (https://rgc-covid19.regeneron.com/terms-of-use).

## Acknowledgements

This research has been conducted using the UK Biobank Resource (Project 26041). The Penn Medicine BioBank is funded by a gift from the Smilow family, the National Center for Advancing Translational Sciences of the National Institutes of Health under CTSA Award Number UL1TR001878, and the Perelman School of Medicine at the University of Pennsylvania. We want to acknowledge the participants and investigators of the FinnGen study. We thank the AncestryDNA customers who voluntarily contributed information in the COVID-19 survey.

## SUPPLEMENTARY FIGURES

Provided in a separate file.

**Supplementary Figure 1.**
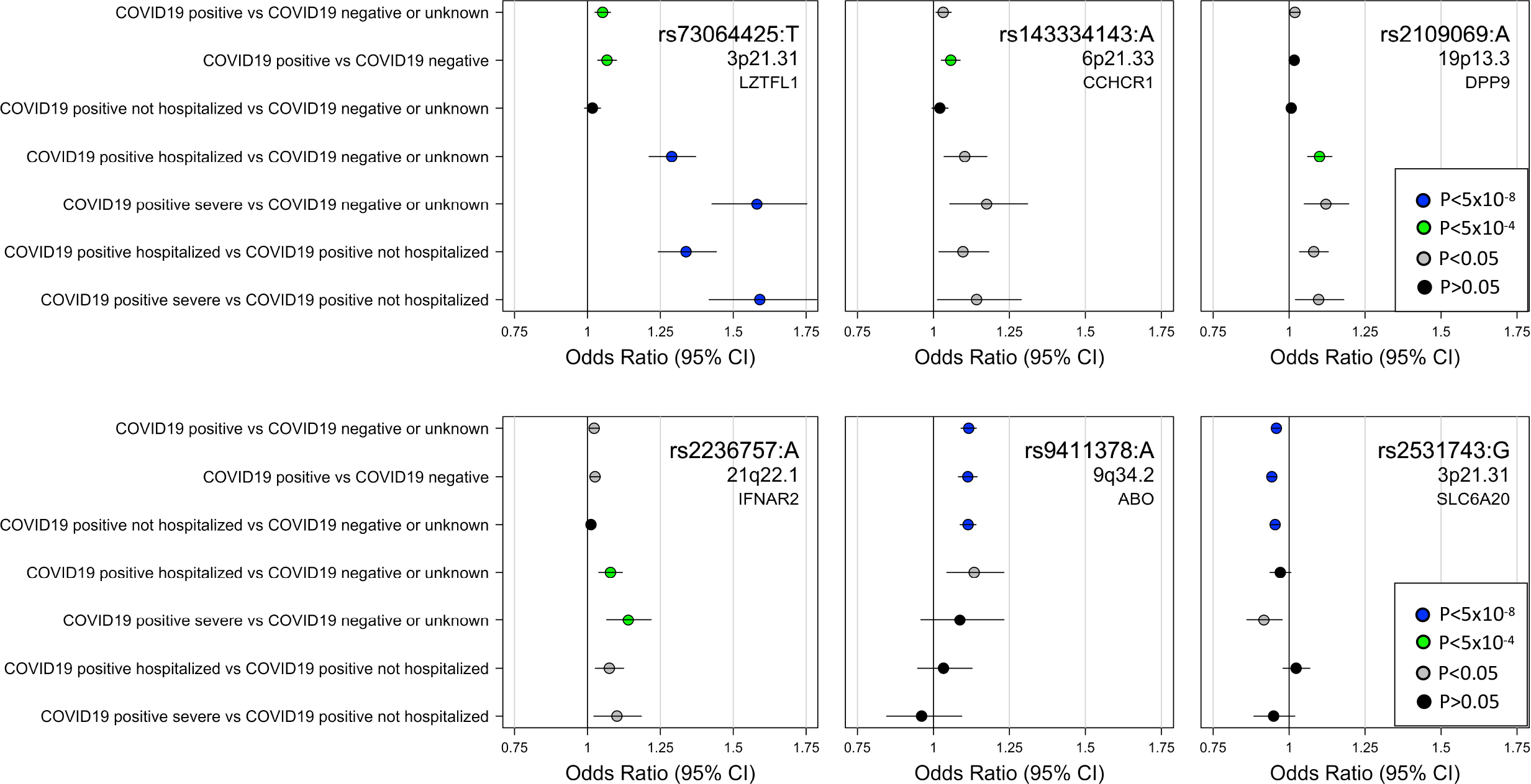
Comparison of effect sizes across COVID-19 risk and severity outcomes for six previously reported novel risk variants that validated in this study. Six variants were reported to associatd with risk of COVID-19 in previous studies and replicated in our analysis. Of these, four variants also associated with disease severity among COVID-19 cases (in/near *LZTFL1*, *CCHCR1*, *DPP9* and *IFNAR2*), whereas two variants did not (in *ABO* and *SLC6A20*). Error bars represent 95% confidence intervals.

**Supplementary Figure 2.**
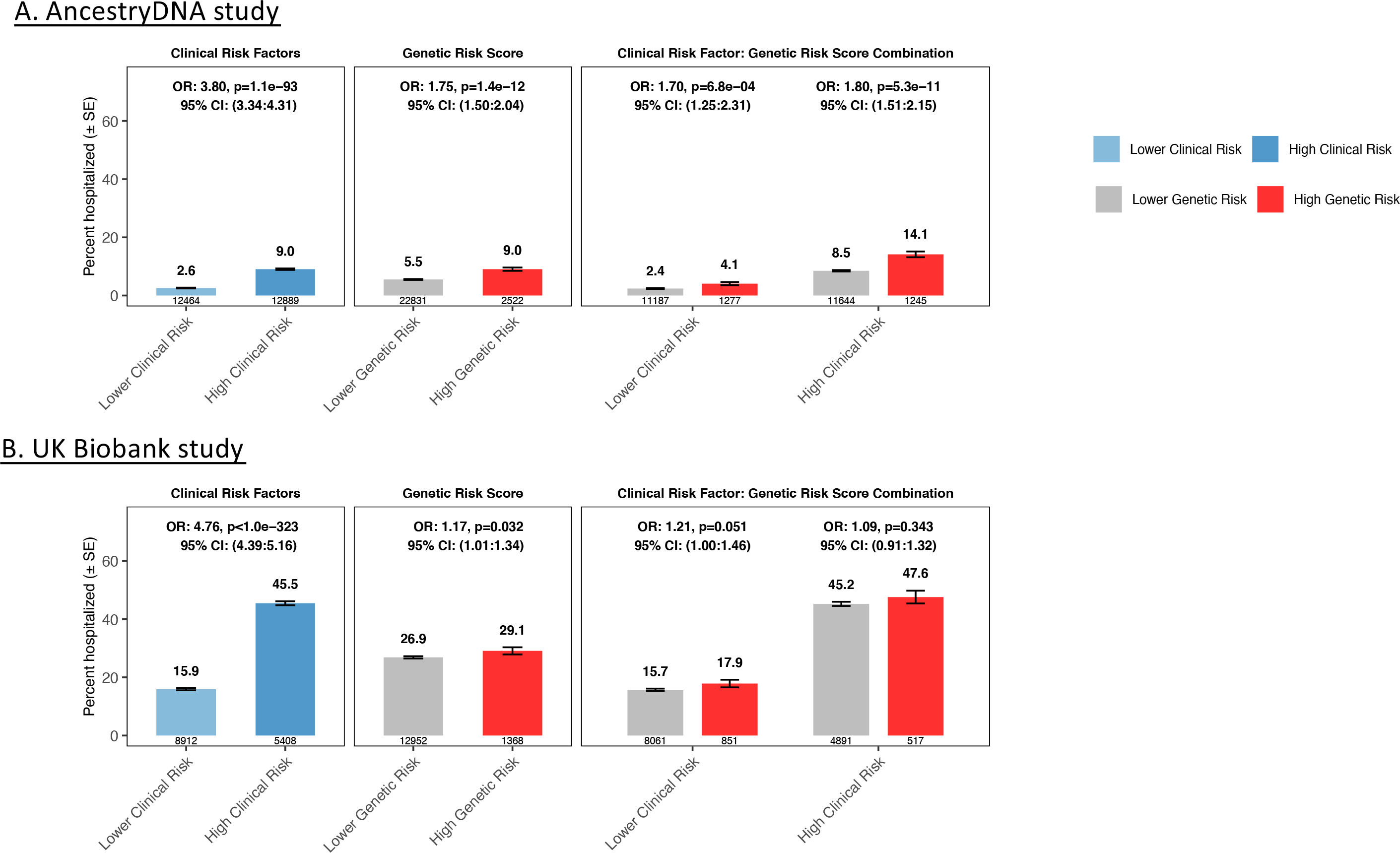
Association between a 6-SNP genetic risk score (GRS) and risk of hospitalization among COVID-19 cases of European ancestry after stratifying by the presence of clinical risk factors. (**A**) Rate of hospitalization in the AncestryDNA study (25,353 COVID-19 cases, including 1,484 hospitalized). (B) Rate of hospitalization in the UK Biobank study (14,320 COVID-19 cases, including 3,878 hospitalized). High genetic risk (red bars): top 10% of the GRS. Low genetic risk (grey bars): bottom 90% of the GRS (*i.e.* all other COVID-19 cases).

**Supplementary Figure 3.**
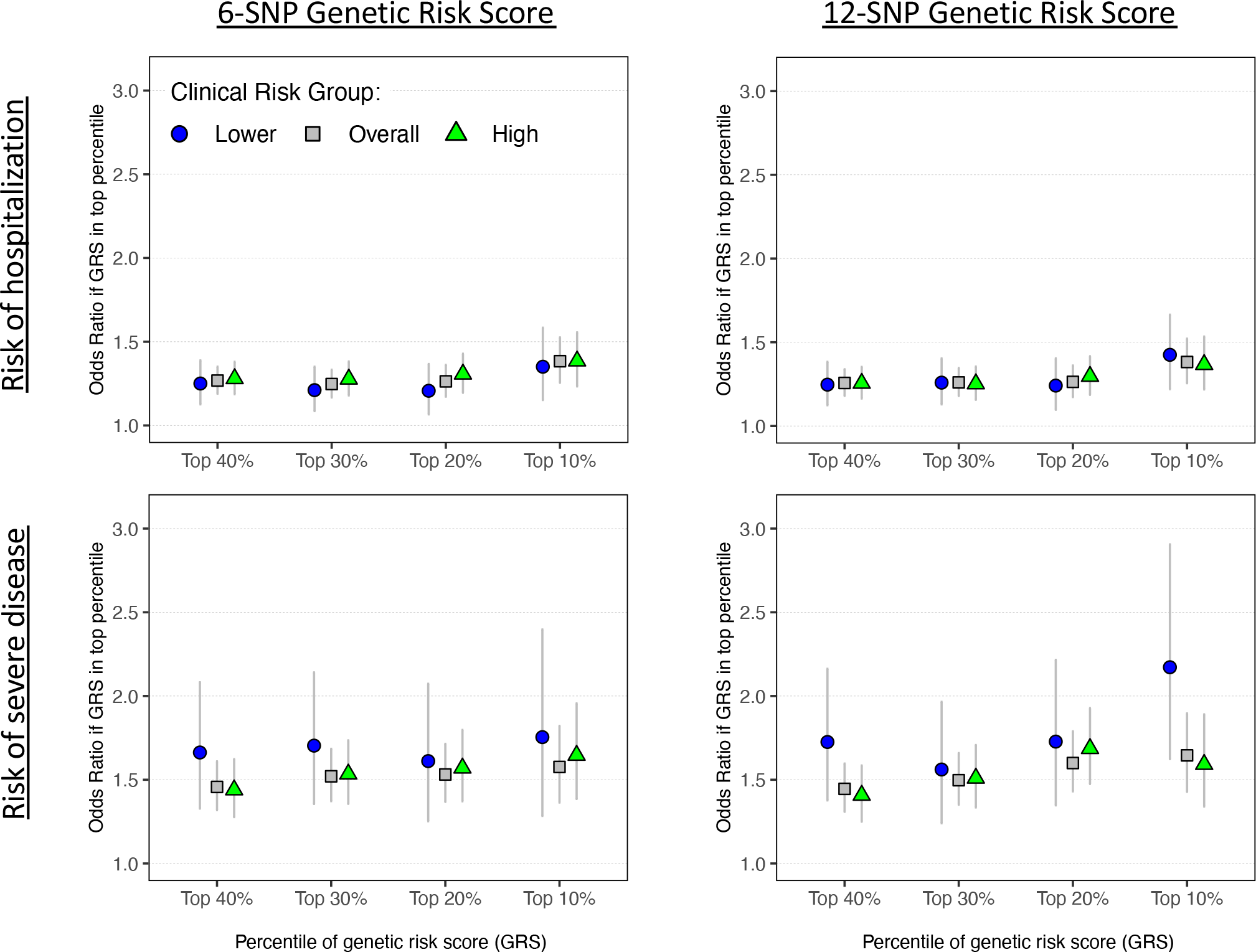
Association between a 6- and 12-SNP genetic risk score (GRS) and risk of hospitalization and severe disease among COVID-19 cases of European ancestry. To evaluate if the association between the GRS and worse disease outcomes was dependent on the list of variants selected for analysis, we compared results between GRS calculated using different sets of variants. We considered a GRS calculated using: (i) the six variants that were reported in previous GWAS of COVID-19 and that we validated the published association and further showed that they were associated with risk of hospitalization or severe disease among COVID-19 cases (in/near *LZTFL1*, MHC, *DPP9*, *IFNAR2*, *RPL24* and *FOXP4*; see Supplementary **Figure 1**); or (ii) all 12 variants reported in previous GWAS of COVID-19 (in/near *LZTFL1* [two variants], MHC, *ABO*, *OAS3*, *DPP9*, *RAVER1*, *IFNAR2*; and four novel risk variants discovered by the HGI in/near *RPL24, DNAH5, FOXP4* and *PLEKHA4*; see **Supplementary Tables 5 and 15**). Analyses were performed separately in the UK Biobank, AncestryDNA and GHS studies (risk of hospitalization only) after stratifying COVID-19 cases by the presence of clinical risk factors, considering individuals with lower clinical risk (blue circles), high clinical risk (green triangles) or all individuals (grey squares). Association results were then meta-analyzed across studies.

**Supplementary Figure 4.**
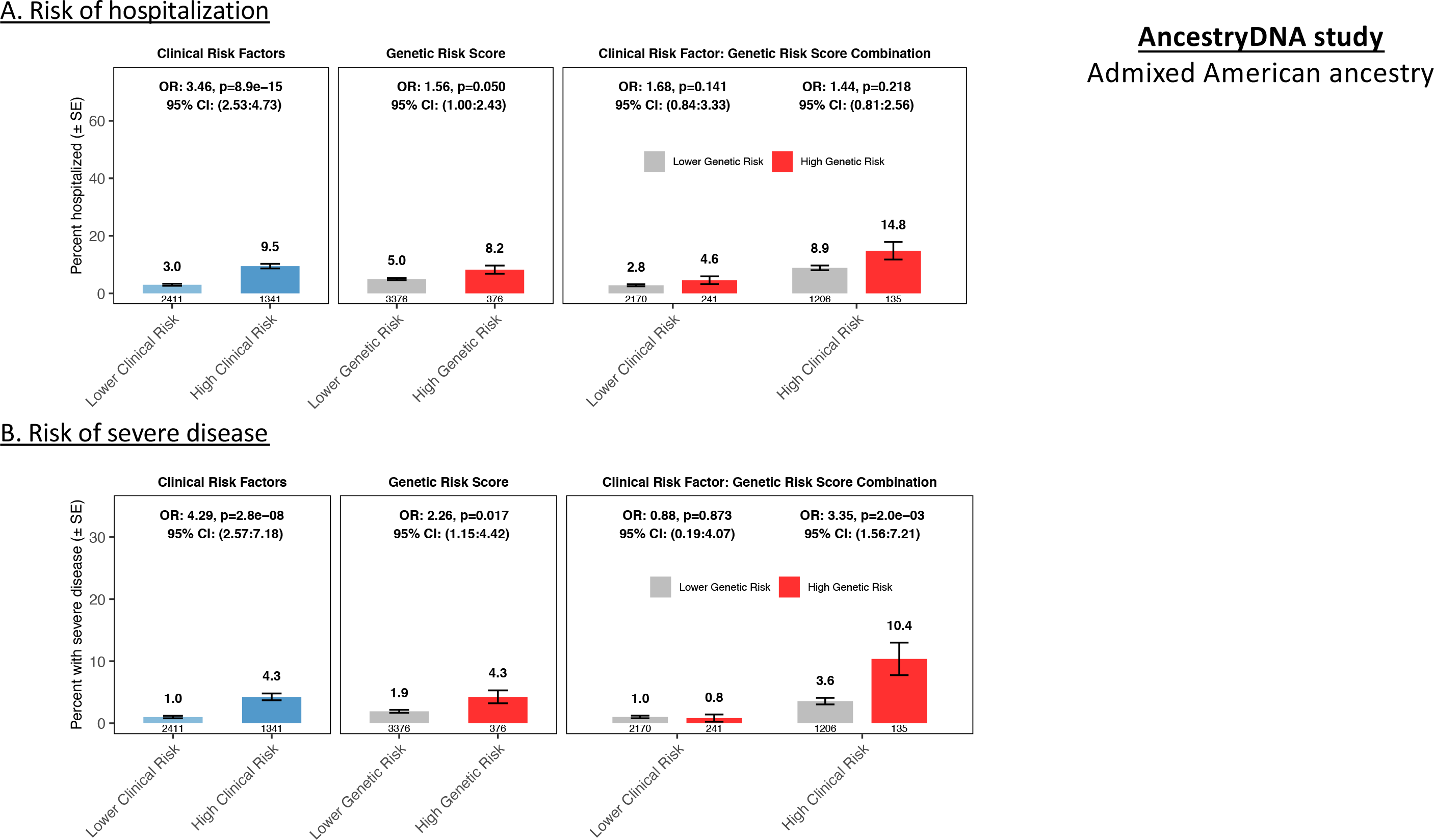
Association between a six-SNP genetic risk score (GRS) and risk of hospitalization (A) and (B) severe disease among COVID-19 cases of Admixed American ancestry.

**Supplementary Figure 5.**
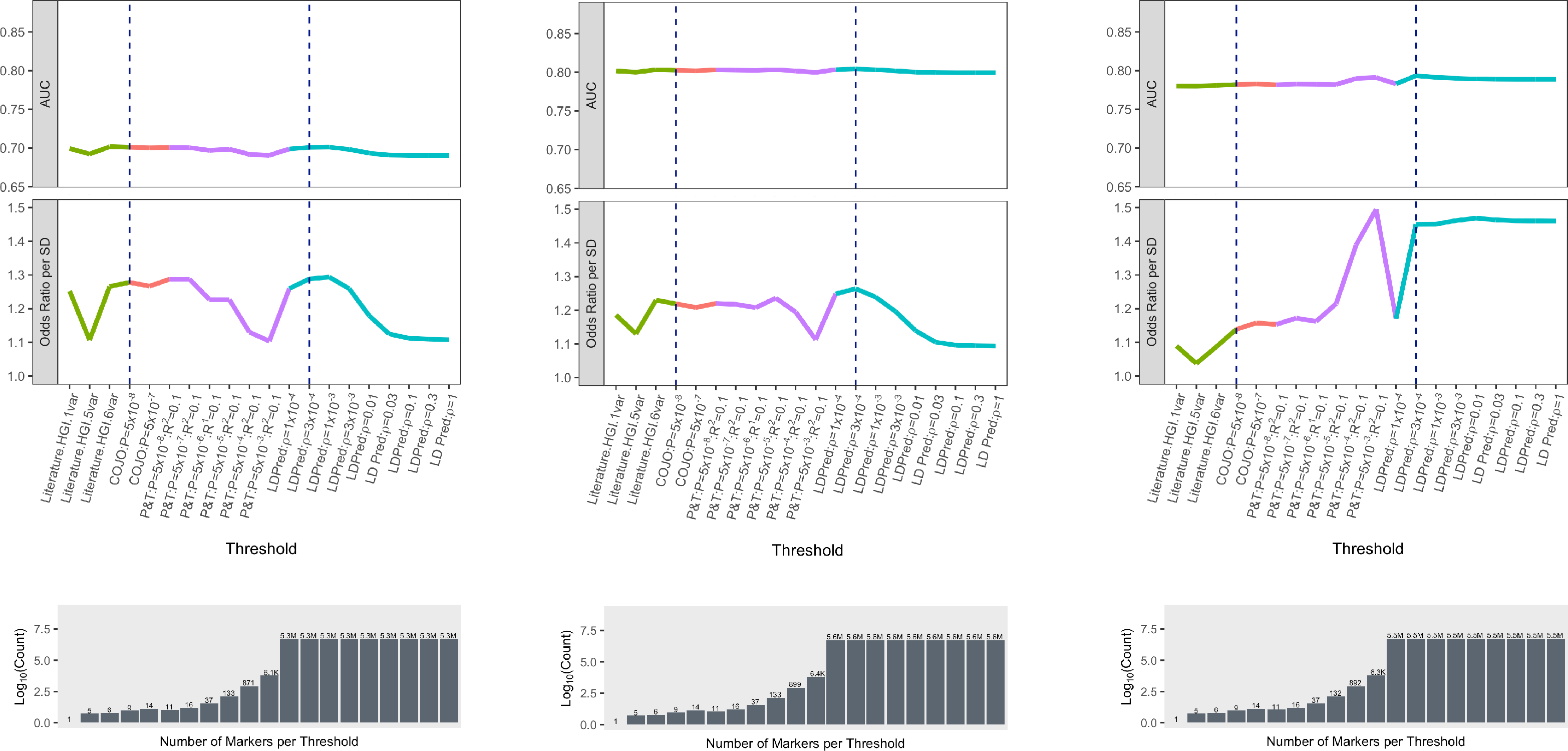
Association between risk of severe disease among COVID-19 cases of European ancestry and genetic risk scores determined based on different criteria. We compared GRS based on variants (i) that were reported in the literature and validated in this study (Literature.HGI.1var: rs73064425 in LZTFL1; Literature.HGI.5var: variants from our 6-SNP model, with the exception of rs73064425 in LZTFL1; Literature.HGI.6var: all six variants from our 6-SNP model); (ii) obtained through pruning and thresholding applied to results from the risk of infection phenotype reported by the HGI, using different association P-value and LD *r*^2^ thresholds; (iii) the LDpred approach ^39^ applied to risk of infection reported by the HGI, considering different ϱ parameters.

**Supplementary Figure 6.**
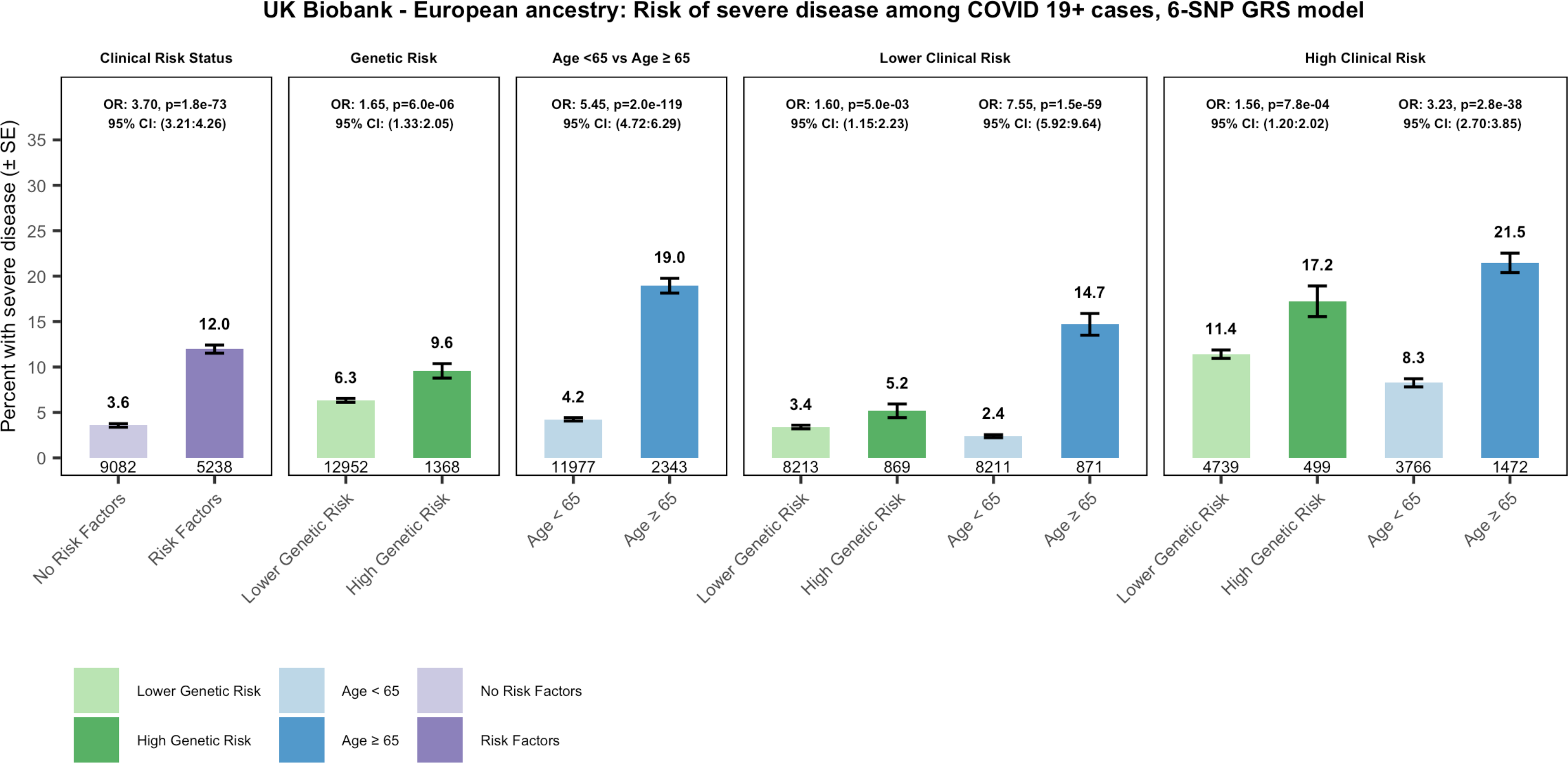
Association between risk of severe disease among COVID-19 cases of European ancestry from the UK Biobank study and combination of clinical risk factors, genetic risk (6-SNP GRS model) and age.

## SUPPLEMENTARY TABLES

Provided in a separate file.

**Supplementary Table 1.**
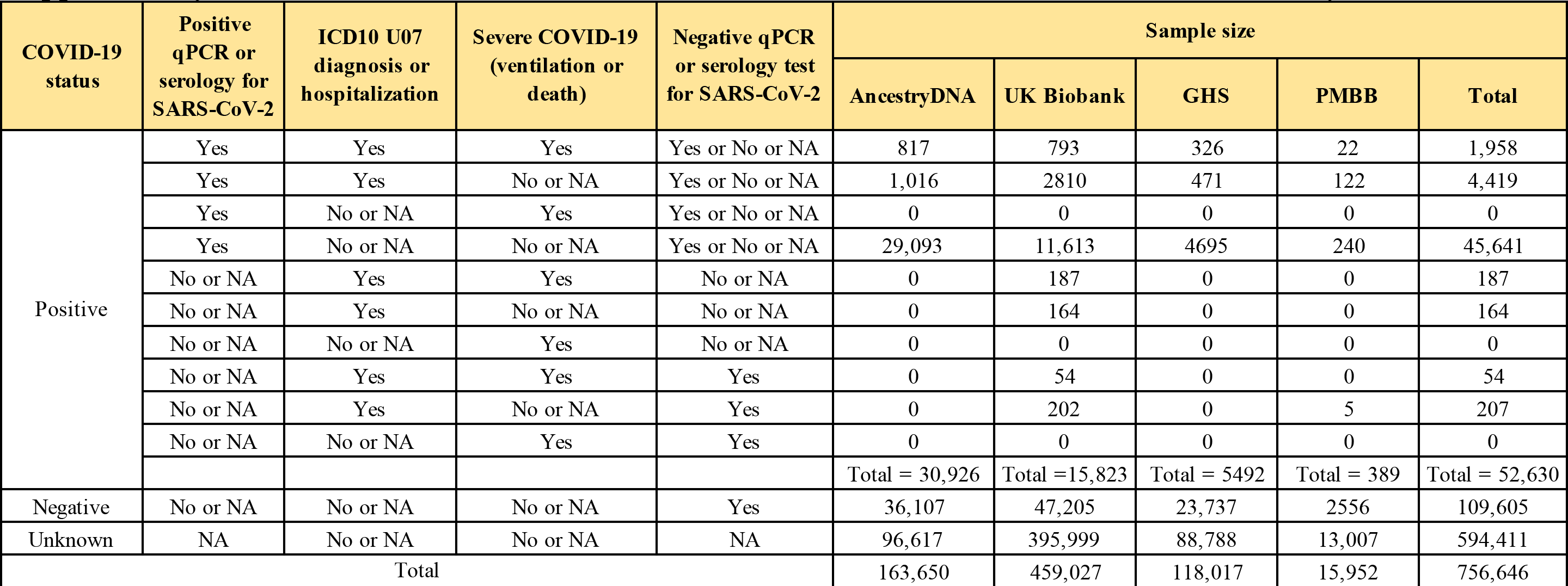
Breakdown of COVID-19 status across the four studies included in the analysis.

**Supplementary Table 2.**
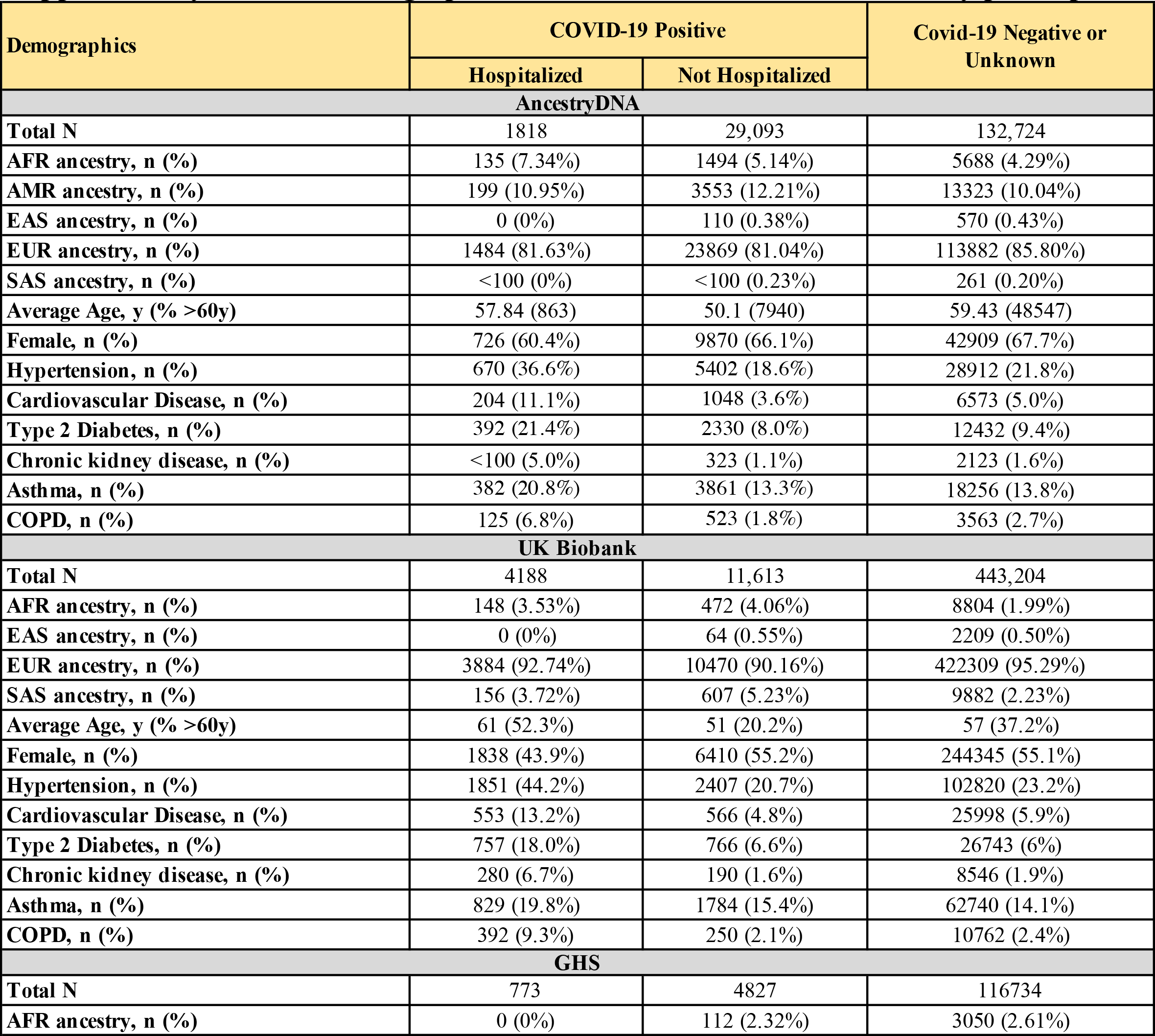
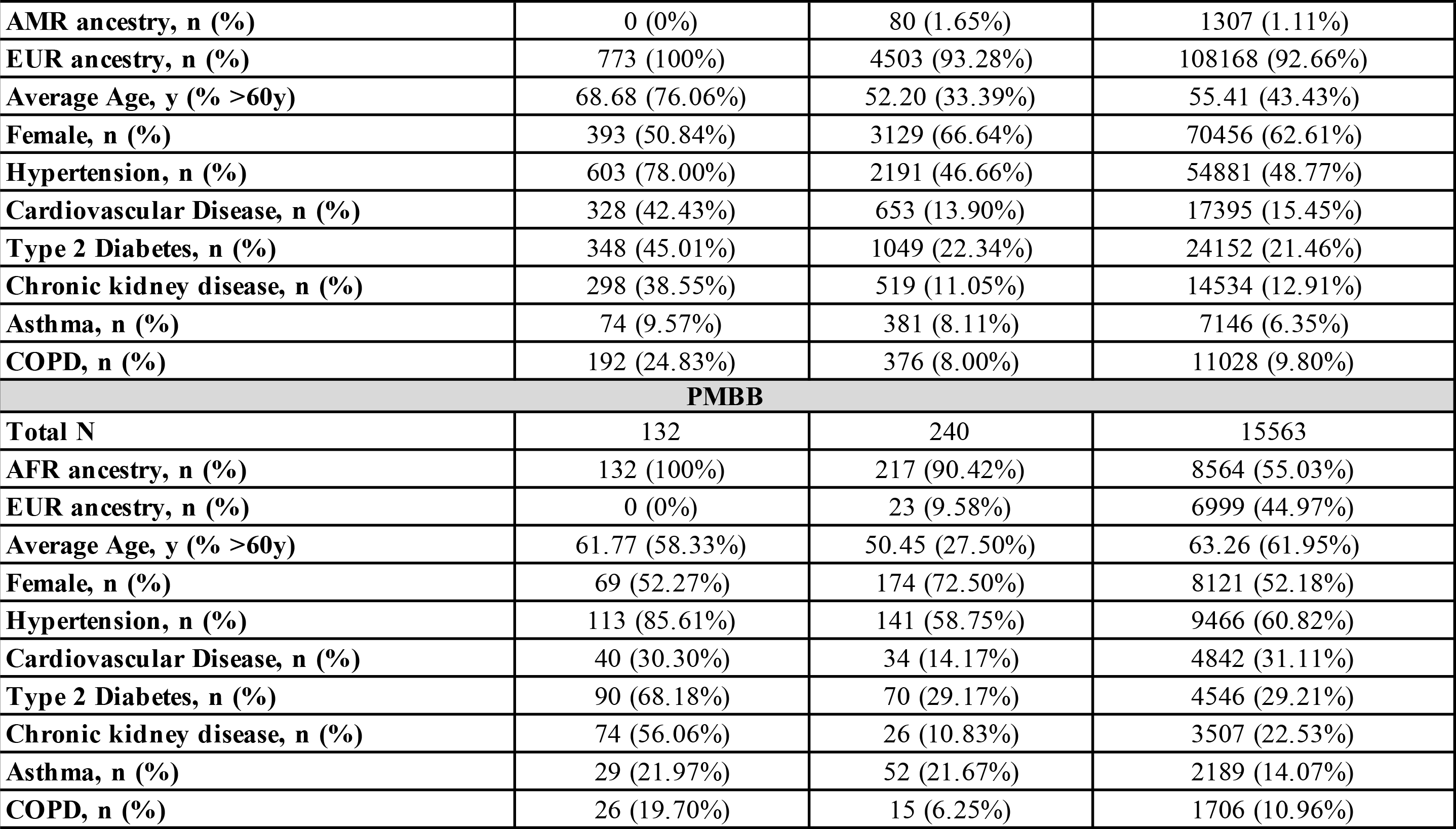
Demographics and clinical characteristics of study participants.

**Supplementary Table 3.**
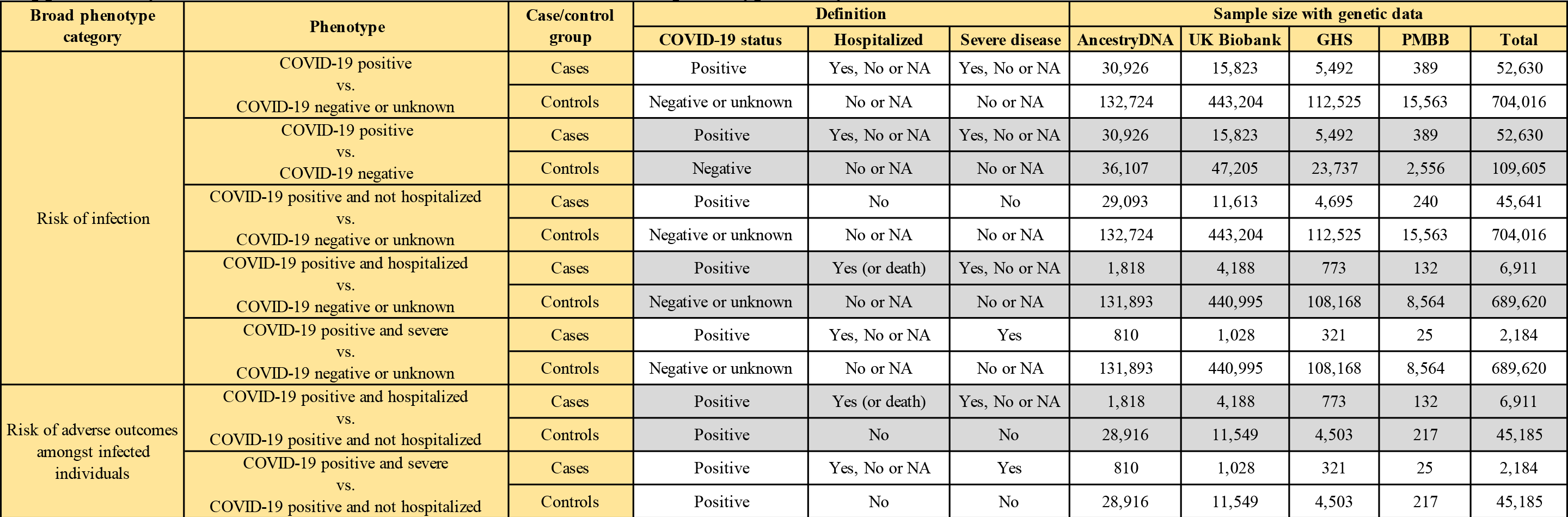
Definitions used for the seven COVID-19 phenotypes analyzed.

**Supplementary Table 4.**
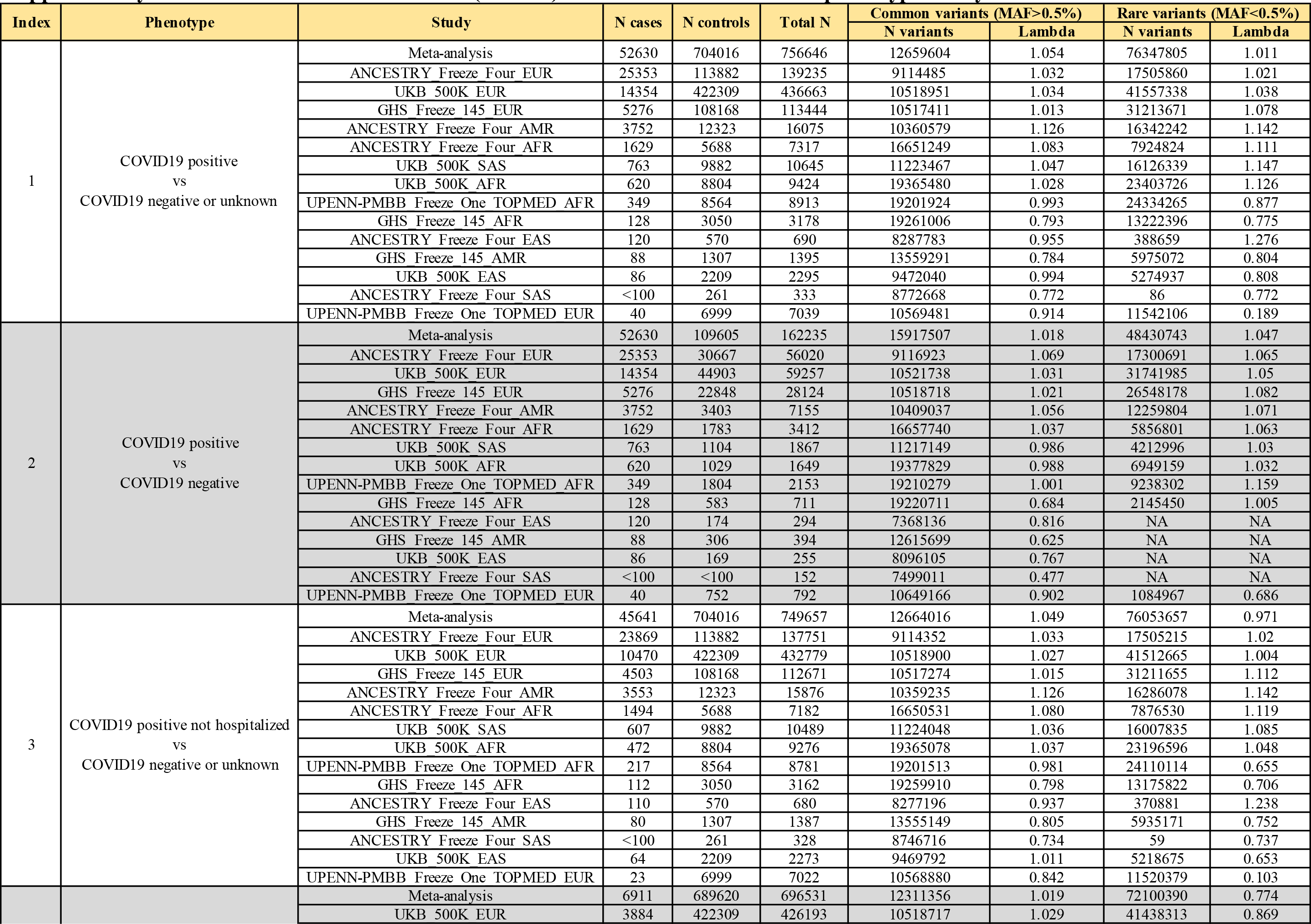
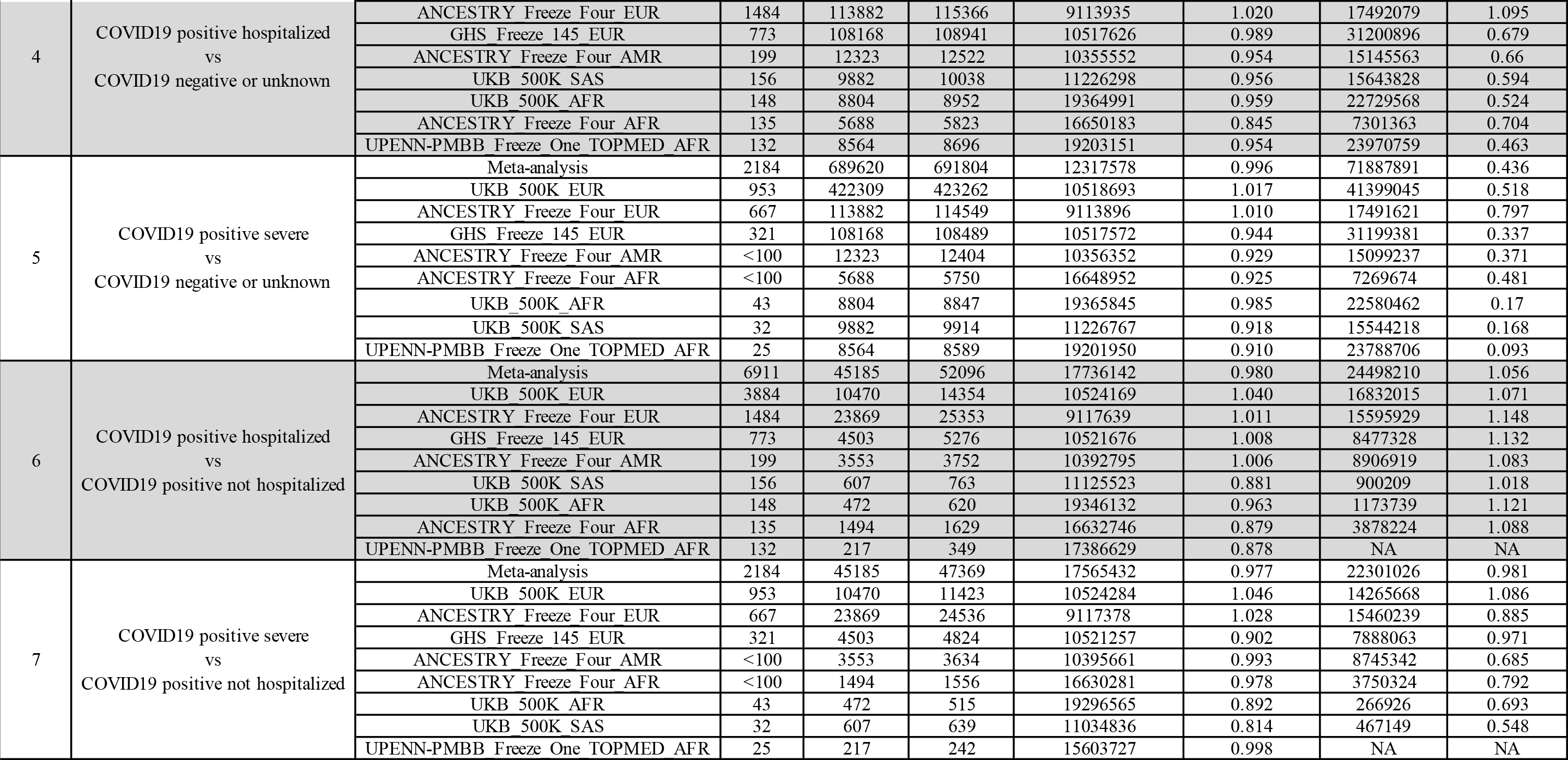
Genomic inflation factor (lambda) across the seven COVID-19 phenotypes analyzed.

**Supplementary Table 5.** Eight variants associated with COVID-19 susceptibility in previous GWAS.

| Locus index | Independent signal index | Reference (PMID) | First author | Phenotype | rs ID | hg38 | Nearest gene | Effect allele | Locus | Odds Ratio | LCI | UCI | P-value | N cases | N controls | Top independent association in locus |
| --- | --- | --- | --- | --- | --- | --- | --- | --- | --- | --- | --- | --- | --- | --- | --- | --- |
| 1 | 1 | 33307546 | Pairo-Castineira | covid19 severe vs negative or unknown | rs73064425 | 3:45859597:C:T | *LZTFL1* | T | 3p21.31 | 2.14 | 1.88 | 2.45 | 4.77E-30 | 1676 | 8380 | 1 |
| 1 | 2 | 33888907 | Shelton | covid test positive vs negative | rs2531743 | 3:45796808:G:A | *SLC6A20* | G | 3p21.31 | 0.917 | 0.89 | 0.94 | 7.60E-10 | 12972 | 101268 | 1 |
| 2 | 1 | 33307546 | Pairo-Castineira | covid19 severe vs negative or unknown | rs143334143 | 6:31153649:G:A | *CCHCR1* | A | 6p21.33 | 1.85 | 1.61 | 2.13 | 8.82E-18 | 1676 | 8380 | 1 |
| 3 | 1 | 33888907 | Shelton | covid test positive vs negative | rs9411378 | 9:133270015:A:C | *ABO* | A | 9q34.2 | 1.17 | 1.12 | 1.20 | 5.30E-20 | 12972 | 101268 | 1 |
| 4 | 1 | 33307546 | Pairo-Castineira | covid19 severe vs negative or unknown | rs10735079 | 12:112942203:G:A | *OAS3* | A | 12q24.13 | 1.29 | 1.18 | 1.42 | 1.65E-08 | 1676 | 8380 | 1 |
| 5 | 1 | 33307546 | Pairo-Castineira | covid19 severe vs negative or unknown | rs2109069 | 19:4719431:G:A | *DPP9* | A | 19p13.3 | 1.36 | 1.25 | 1.48 | 3.98E-12 | 1676 | 8380 | 1 |
| 6 | 1 | 33307546 | Pairo-Castineira | covid19 severe vs negative or unknown | rs74956615 | 19:10317045:T:A | *RAVER1* | A | 19p13.2 | 1.59 | 1.35 | 1.87 | 2.31E-08 | 1676 | 8380 | 1 |
| 7 | 1 | 33307546 | Pairo-Castineira | covid19 severe vs negative or unknown | rs2236757 | 21:33252612:A:G | *IFNAR2* | A | 21q22.1 | 1.28 | 1.17 | 1.41 | 5.00E-08 | 1676 | 8380 | 1 |

**Supplementary Table 6.** Sample overlap between our study and those queried to identify variants associated with COVID-19 susceptibility in previous GWAS.

| First author | Reference (PMID) | N cases | N controls | Sample overlap with our study |
| --- | --- | --- | --- | --- |
| ***Three studies with minimal or no sample overlap with our study*** |  |  |  |  |
| *(eight variants selected for replication with five susceptibility phenotypes: in/near LZTFL1, SLC6A20, MHC, ABO, OAS3, DPP9, RAVR1 and IFNAR2)* |  |  |  |  |
| Ellinghaus | 32558485 | 1,610 | 2,205 | No known overlap |
| Pairo-Castineira | 33307546 | 1,676 | 8,380 | ~1% of samples from our analysis of UKB were included as controls in this study |
| Shelton | 33888907 | 15,434 | 1,035,598 | No known overlap |
| ***One study with substantial sample overlap with our study*** |  |  |  |  |
| *(Four variants associated with reported infection [phenotype "C2"] selected for look-up in our independent analysis of severity among cases: in/near RPL24, DNAH5, FOXP4 and PLEKHA4)* |  |  |  |  |
| HGI (freeze 5) | NA | 49,562 | 1.7 million | ~100% of samples from UKB, GHS and UPENN, and ~60% from AncestryDNA contributed to HGI, but using an earlier phenotype freeze. |

**Supplementary Table 7.** Comparison of effect sizes between ancestries for the novel ACE2 risk variant.

| Locus,<br>nearest gene | rs ID | hg38 | Effect allele | Phenotype | Ancestry | Odds Ratio (95% CI) | P-value | N cases with 0\|1\|2<br>copies of the effect<br>allele | N controls with 0\|1\|2<br>copies of the effect<br>allele | Effect allele<br>frequency | Cross-ancestry<br>heterogeneity test P-value |
| --- | --- | --- | --- | --- | --- | --- | --- | --- | --- | --- | --- |
| Xp22.2<br>ACE2 | rs190509934 | 23:15602217:T:C | C | COVID19 positive vs<br>COVID19 negative or<br>unknown | AFR | 0.578[0.373,0.894] | 0.014 | 2713\|12\|1 | 25905\|146\|55 | 0.005 | 0.676 |
|  |  |  |  |  | AMR | 0.792[0.431,1.456] | 0.45 | 3830\|10\|0 | 13577\|44\|9 | 0.002 |  |
|  |  |  |  |  | EUR | 0.606[0.516,0.711] | 8.80E-10 | 44930\|87\|6 | 648877\|1835\|646 | 0.002 |  |
|  |  |  |  |  | SAS | 0.500[0.323,0.773] | 0.0018 | 821\|13\|1 | 9815\|226\|102 | 0.020 |  |

**Supplementary Table 8.**
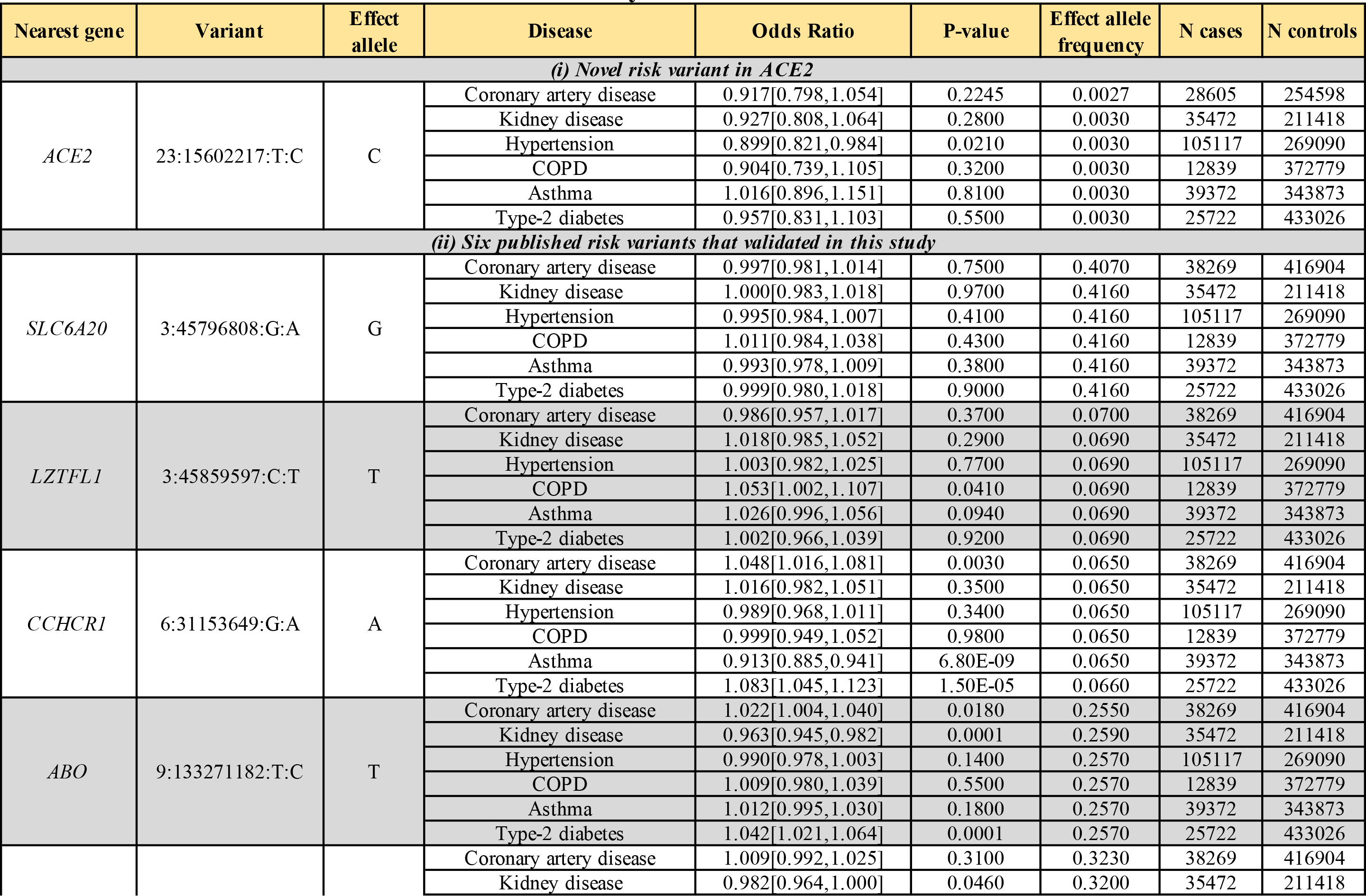
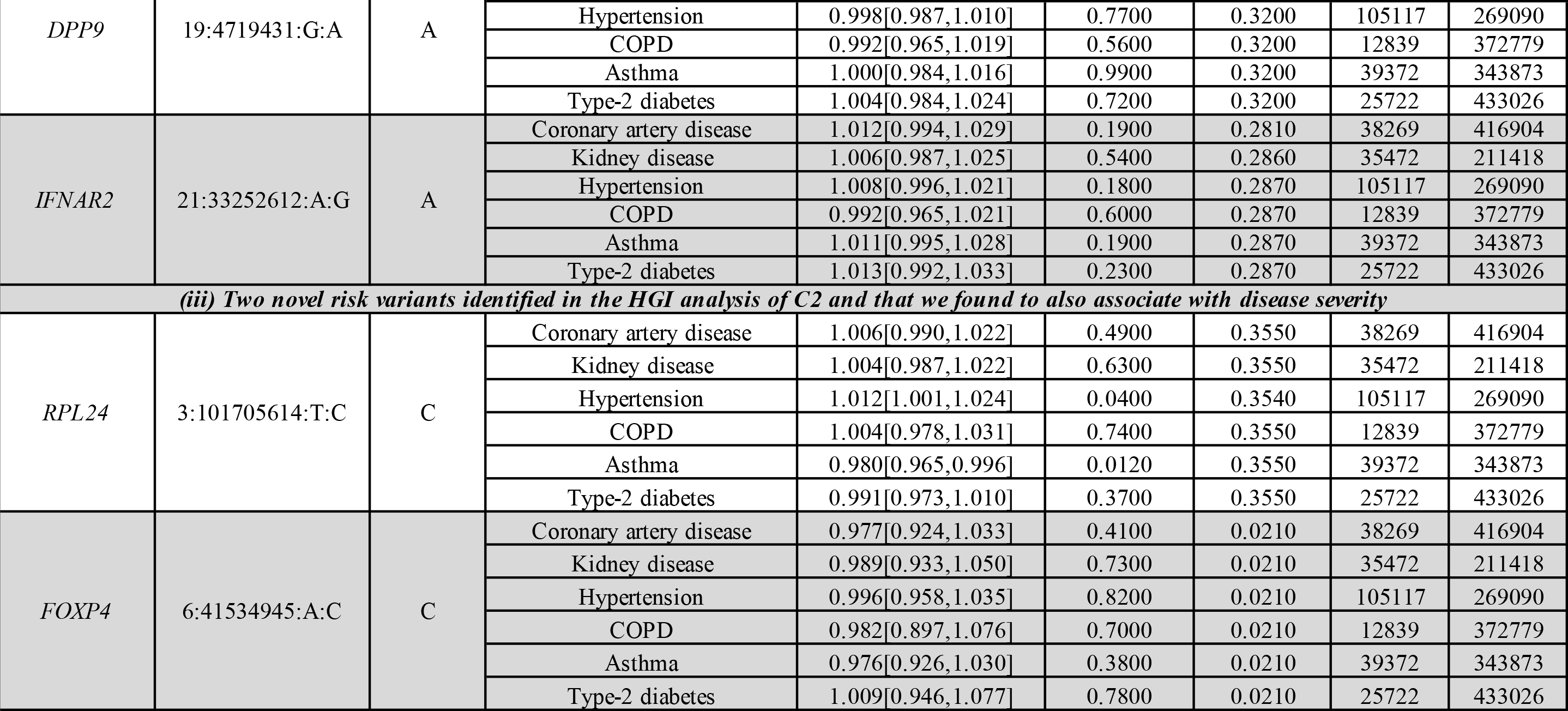
Association results in the UK Biobank study between six established clinical risk factors for COVID-19 and (i) the novel risk variant in ACE2; (ii) the six published risk variants for COVID-19 that validated in this study; and (iii) the two novel risk variants identified in the HGI analysis of C2 and that we found to also associate with disease severity.

**Supplementary Table 9.** Association between two severity phenotypes and a novel risk variant for COVID-19 near ACE2 identified in this study.

| Locus, nearest gene | rs ID | hg38 | Phenotype | Effect allele | Odds Ratio* [95% CI] | P-value | N cases with 0/1/2 copies of the effect allele | N controls with 0/1/2 copies of the effect allele | Effect allele frequency | N cases | N controls | Cross-study heterogeneity test P-value |
| --- | --- | --- | --- | --- | --- | --- | --- | --- | --- | --- | --- | --- |
| Xp22.2<br>ACE2 | rs190509934 | 23:15602217:T:C | COVID19 positive hospitalized vs COVID19 positive not hospitalized | C | 0.694[0.419,1.150] | 1.57E-01 | 6766\|13\|0 | 44854\|106\|8 | 0.001 | 6779 | 44968 | 0.222 |
|  |  |  | COVID19 positive severe vs COVID19 positive not hospitalized | C | 0.544[0.210,1.408] | 2.10E-01 | 2157\|2\|0 | 44854\|106\|8 | 0.001 | 2159 | 44968 | 0.916 |

\* Estimated based on trans-ancestry inverse-variance meta-analysis across UKB, AncestryDNA, GHS and UPENN-PMBB.

**Supplementary Table 10.**
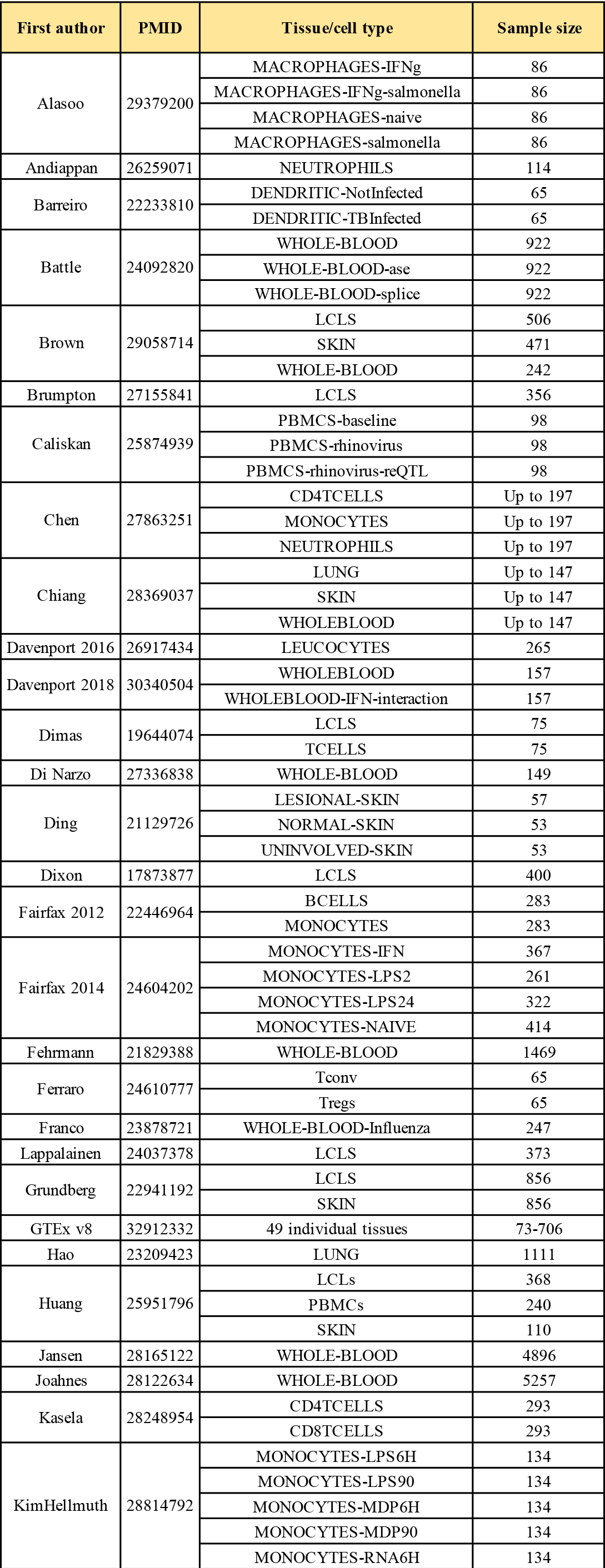
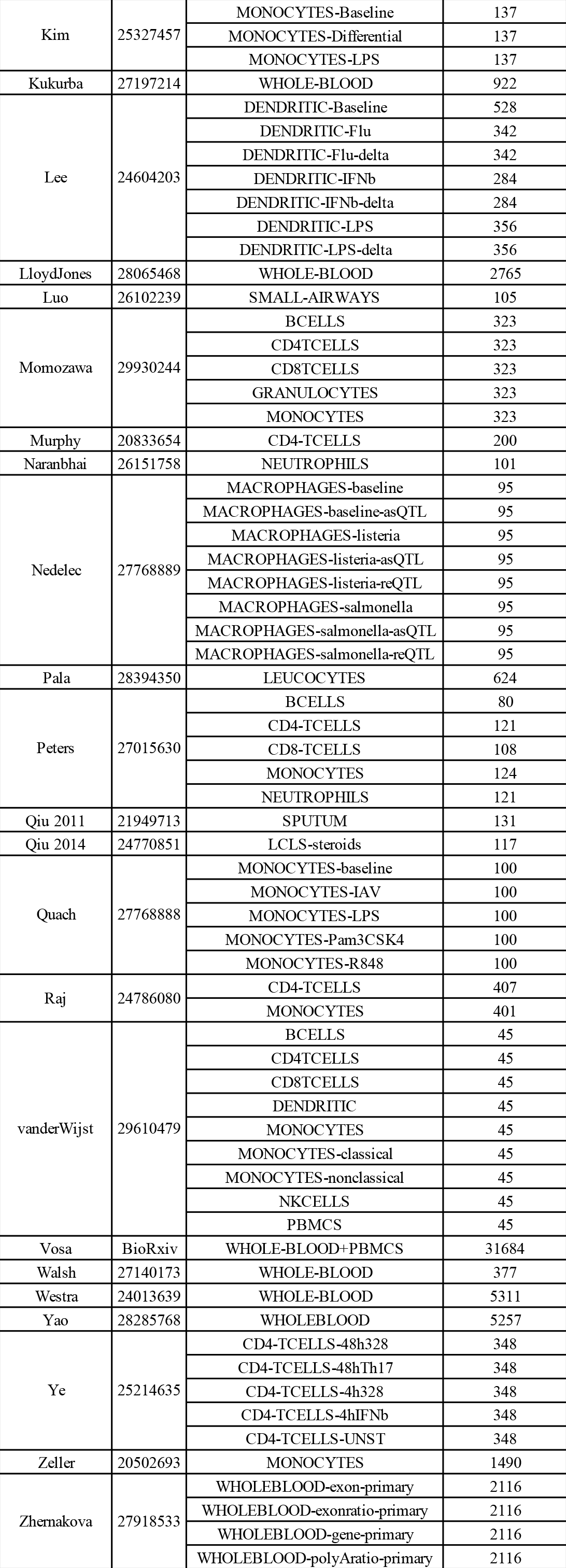
Published gene expression datasets used to identify sentinel expression quantitative trait loci (eQTL) that co-localized (LD r2>0.8) with sentinel GWAS variants.

**Supplementary Table 11.** Association between systolic and diastolic blood pressure in Europeans of the UK Biobank study and (i) the rare non-coding variant near ACE2 that was found to be associated with COVID-19 risk; and (ii) a burden of ultra-rare coding variants in ACE2 that are predicted to be loss-of-function.

| Variant | Effect allele | Phenotype | Effect[95% CI] | P-value | N individuals with 0\|1\|2 copies of the effect allele | Effect allele frequency |
| --- | --- | --- | --- | --- | --- | --- |
| 23:15602217:T:C | C | Systolic blood pressure | 0.009[-0.022,0.041] | 5.60E-01 | 406539\|1179\|493 | 3.00E-03 |
|  |  | Diastolic blood pressure | 0.017[-0.017,0.051] | 3.30E-01 | 406543\|1179\|493 | 3.00E-03 |
| Burden of predicted loss-of-function (pLOF) variants | Minor allele for any pLOF variant | Systolic blood pressure | 0.219[-0.031,0.469] | 8.60E-02 | 408174\|33\|4 | 5.00E-05 |
|  |  | Diastolic blood pressure | 0.195[-0.070,0.461] | 1.50E-01 | 408178\|33\|4 | 5.00E-05 |

**Supplementary Table 12.**
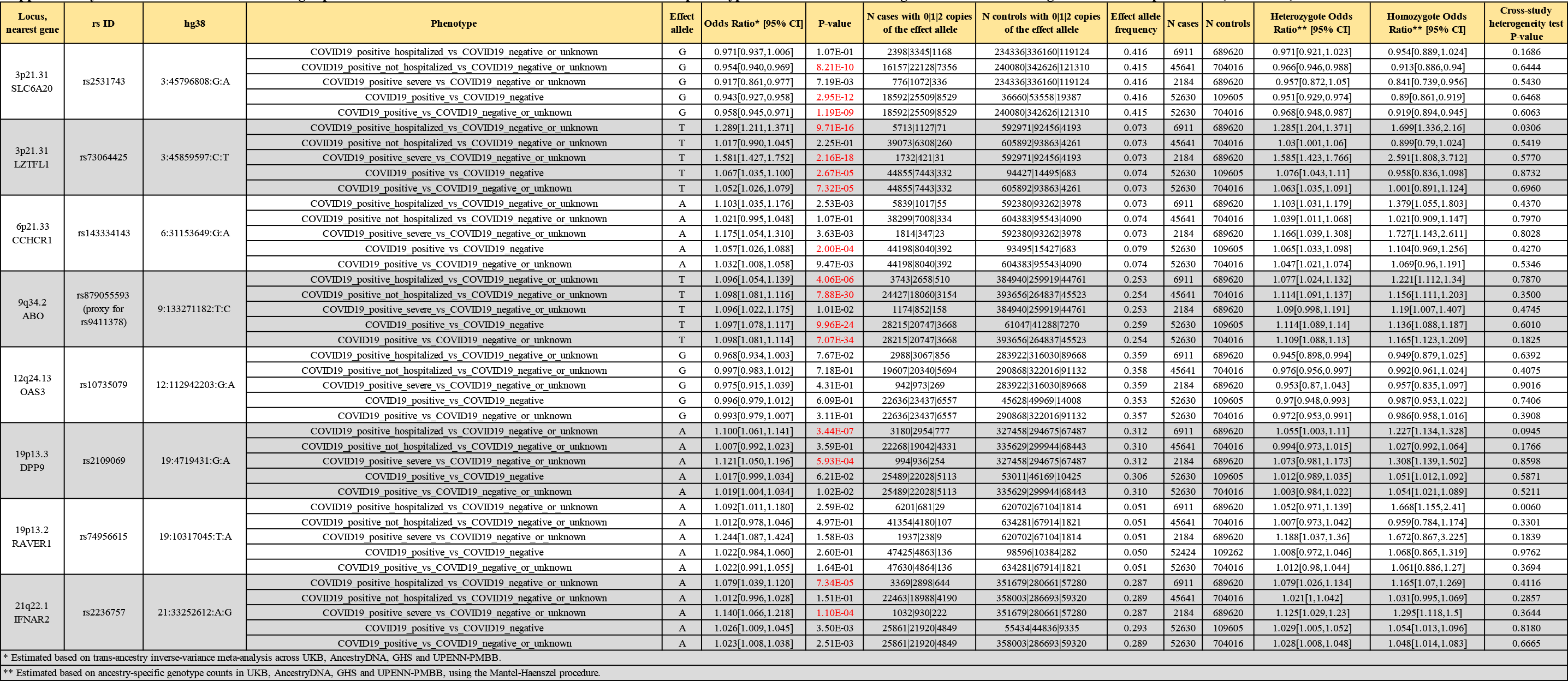
Association between eight published risk variants for COVID-19 and five disease risk phenotypes. P-values in red were significant after correcting for the 40 tests performed (P<0.00125).

**Supplementary Table 13.**
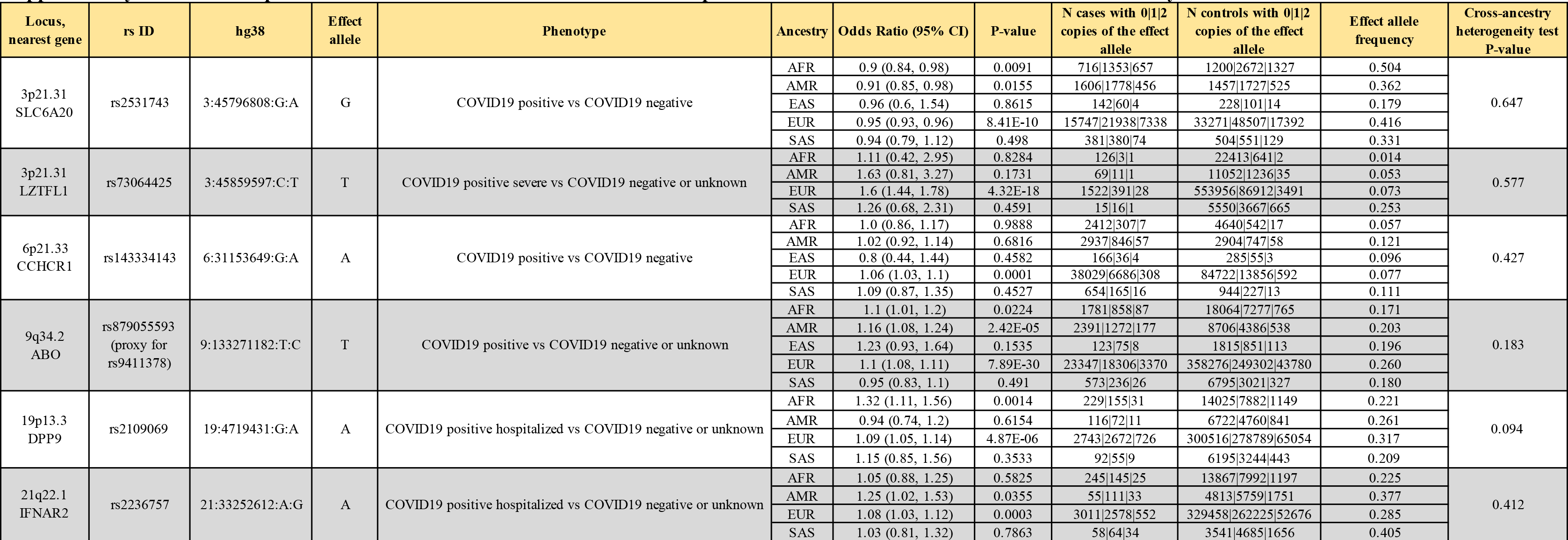
Comparison of effect sizes between ancestries for the six published risk variants that were validated in this study.

**Supplementary Table 14.**
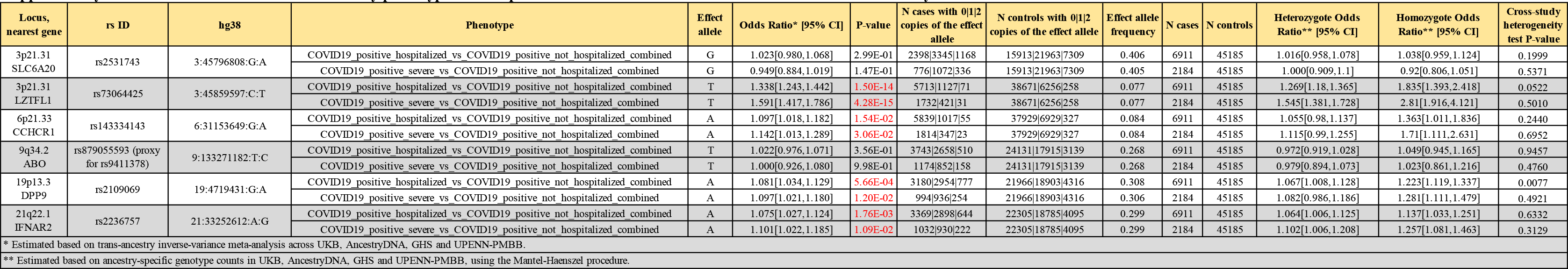
Association between two severity phenotypes and six published risk variants for COVID-19 that validated in this study.

**Supplementary Table 15.**
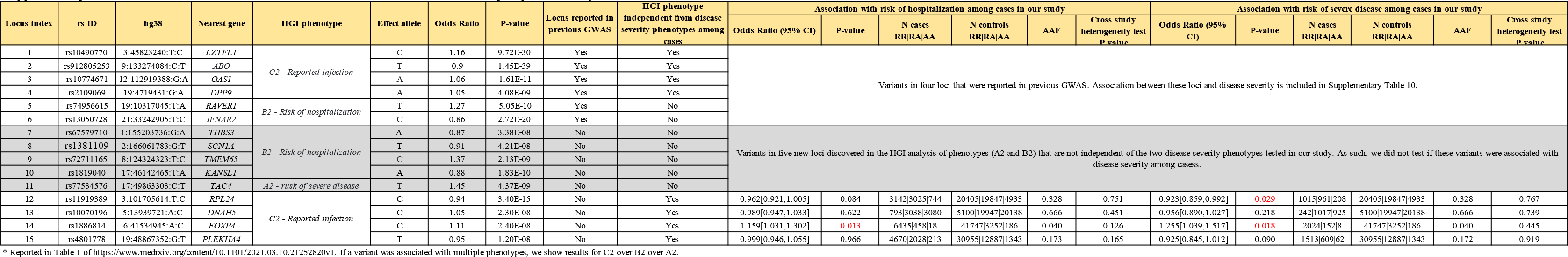
Fifteen variants associated with COVID-19 in the latest meta-analyses performed by the COVID-19 Host Genetics Initiative*.

**Supplementary Table 16.**
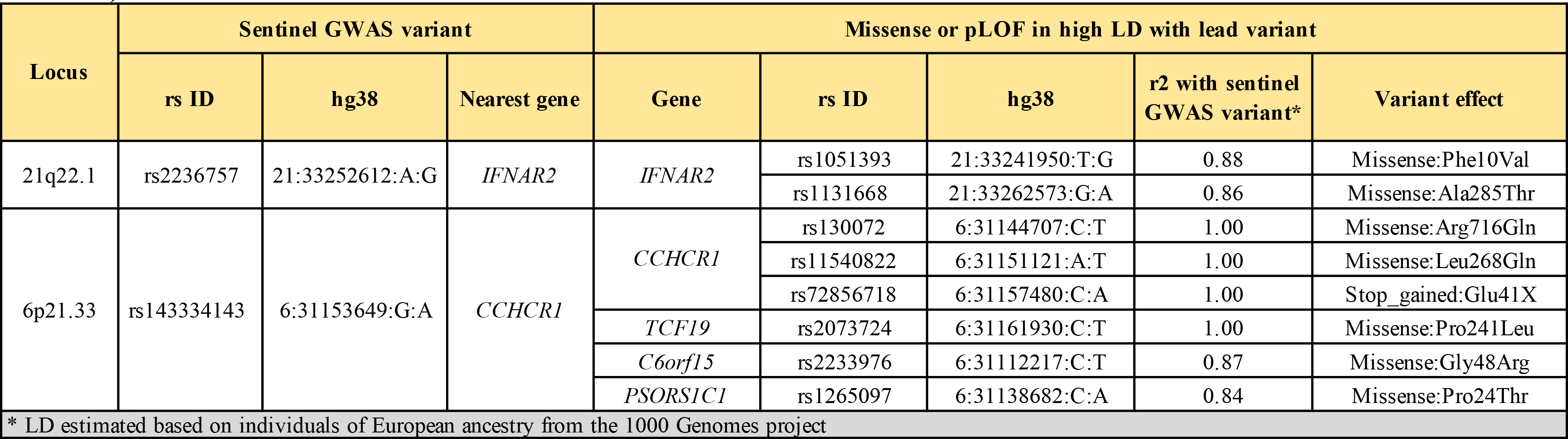
Missense or predicted loss-of-function variants in high linkage disequilibrium (LD, r2>0.80) with sentinel GWAS variants.

**Supplementary Table 17.**
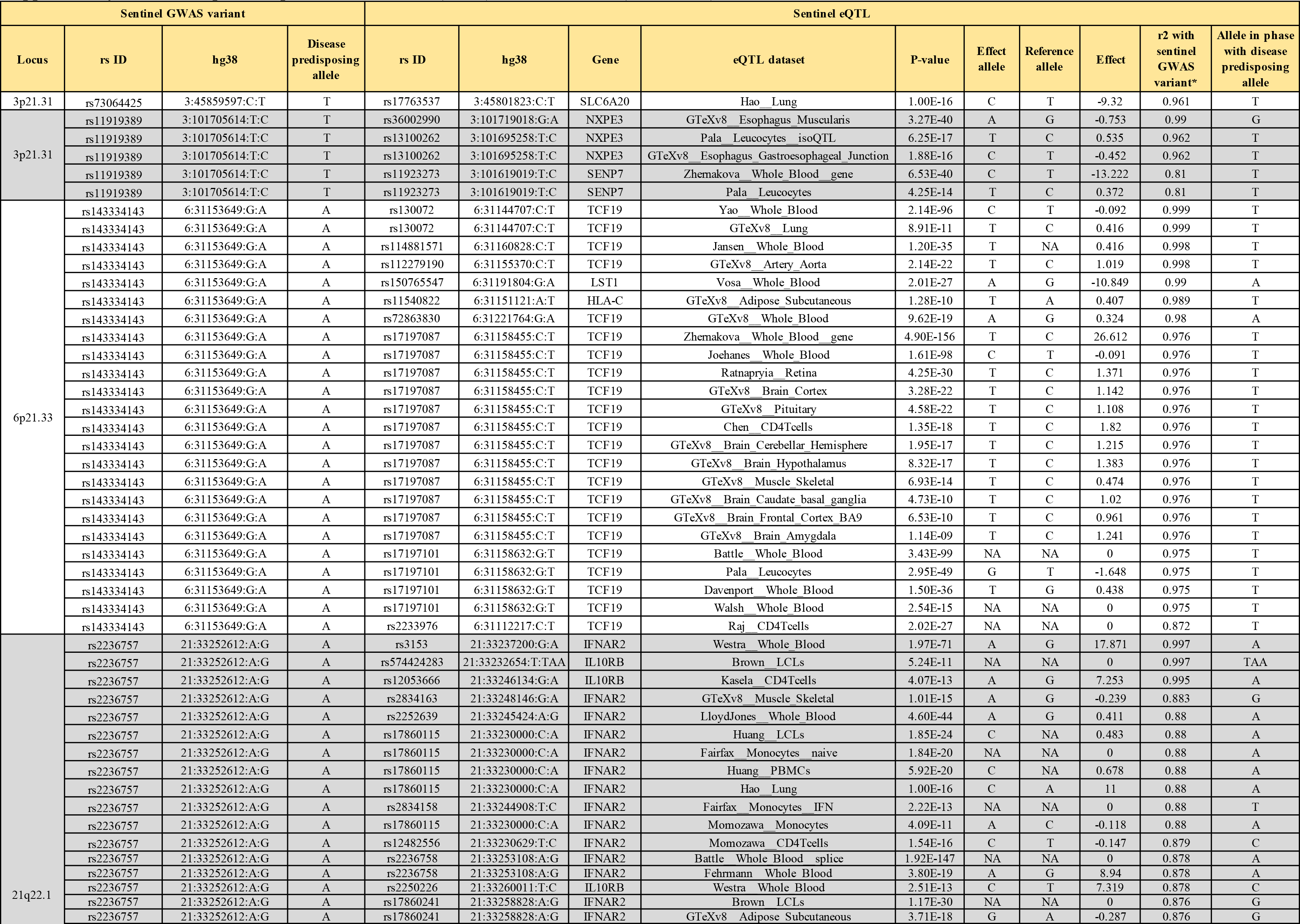
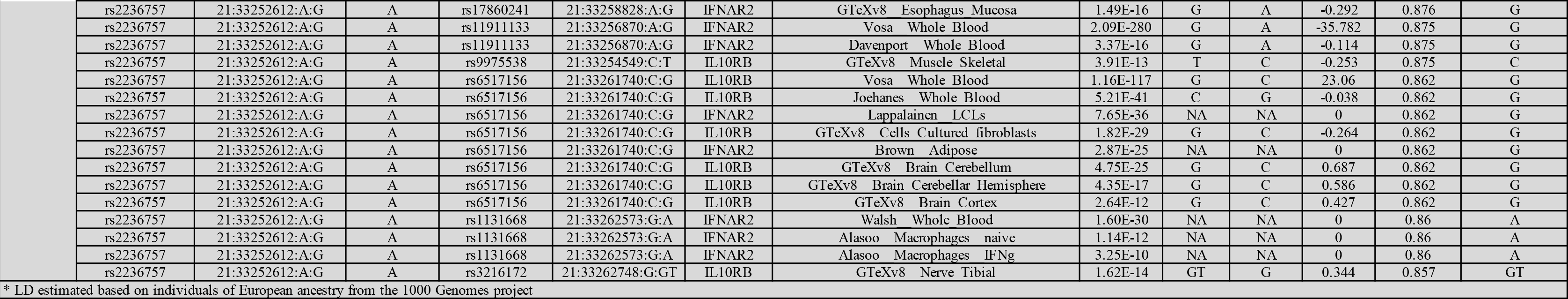
Expression quantitative trait loci (eQTL) that co-localized (LD r2>0.80) with sentinel GWAS variants.

| Sentinel GWAS variant |  |  |  | Sentinel eQTL |  |  |  |  |  |  |  |  |  |
| --- | --- | --- | --- | --- | --- | --- | --- | --- | --- | --- | --- | --- | --- |
| Locus | rs ID | hg38 | Disease predisposing allele | rs ID | hg38 | Gene | eQTL dataset | P-value | Effect allele | Reference allele | Effect | r2 with sentinel GWAS variant* | Allele in phase with disease predisposing allele |
| 3p21.31 | rs73064425 | 3:45859597:C:T | T | rs17763537 | 3:45801823:C:T | SLC6A20 | Hao Lung | 1.00E-16 | C | T | -9.32 | 0.961 | T |
| 3p21.31 | rs11919389 | 3:101705614:T:C | T | rs36002990 | 3:101719018:G:A | NXPE3 | GTeXv8 Esophagus Muscularis | 3.27E-40 | A | G | -0.753 | 0.99 | G |
|  | rs11919389 | 3:101705614:T:C | T | rs13100262 | 3:101695258:T:C | NXPE3 | Pala Leucocytes isoQTL | 6.25E-17 | T | C | 0.535 | 0.962 | T |
|  | rs11919389 | 3:101705614:T:C | T | rs13100262 | 3:101695258:T:C | NXPE3 | GTeXv8 Esophagus Gastroesophageal Junction | 1.88E-16 | C | T | -0.452 | 0.962 | T |
|  | rs11919389 | 3:101705614:T:C | T | rs11923273 | 3:101619019:T:C | SENP7 | Zhemakova Whole Blood gene | 6.53E-40 | C | T | -13.222 | 0.81 | T |
|  | rs11919389 | 3:101705614:T:C | T | rs11923273 | 3:101619019:T:C | SENP7 | Pala Leucocytes | 4.25E-14 | T | C | 0.372 | 0.81 | T |
| 6p21.33 | rs143334143 | 6:31153649:G:A | A | rs130072 | 6:31144707:C:T | TCF19 | Yao Whole Blood | 2.14E-96 | C | T | -0.092 | 0.999 | T |
|  | rs143334143 | 6:31153649:G:A | A | rs130072 | 6:31144707:C:T | TCF19 | GTeXv8 Lung | 8.91E-11 | T | C | 0.416 | 0.999 | T |
|  | rs143334143 | 6:31153649:G:A | A | rs114881571 | 6:31160828:C:T | TCF19 | Jansen Whole Blood | 1.20E-35 | T | NA | 0.416 | 0.998 | T |
|  | rs143334143 | 6:31153649:G:A | A | rs112279190 | 6:31155370:C:T | TCF19 | GTeXv8 Artery Aorta | 2.14E-22 | T | C | 1.019 | 0.998 | T |
|  | rs143334143 | 6:31153649:G:A | A | rs150765547 | 6:31191804:G:A | LST1 | Vosa Whole Blood | 2.01E-27 | A | G | -10.849 | 0.99 | A |
|  | rs143334143 | 6:31153649:G:A | A | rs11540822 | 6:31151121:A:T | HLA-C | GTeXv8 Adipose Subcutaneous | 1.28E-10 | T | A | 0.407 | 0.989 | T |
|  | rs143334143 | 6:31153649:G:A | A | rs72863830 | 6:31221764:G:A | TCF19 | GTeXv8 Whole Blood | 9.62E-19 | A | G | 0.324 | 0.98 | A |
|  | rs143334143 | 6:31153649:G:A | A | rs17197087 | 6:31158455:C:T | TCF19 | Zhemakova Whole Blood gene | 4.90E-156 | T | C | 26.612 | 0.976 | T |
|  | rs143334143 | 6:31153649:G:A | A | rs17197087 | 6:31158455:C:T | TCF19 | Joehanes Whole Blood | 1.61E-98 | C | T | -0.091 | 0.976 | T |
|  | rs143334143 | 6:31153649:G:A | A | rs17197087 | 6:31158455:C:T | TCF19 | Ratnapryia Retina | 4.25E-30 | T | C | 1.371 | 0.976 | T |
|  | rs143334143 | 6:31153649:G:A | A | rs17197087 | 6:31158455:C:T | TCF19 | GTeXv8 Brain Cortex | 3.28E-22 | T | C | 1.142 | 0.976 | T |
|  | rs143334143 | 6:31153649:G:A | A | rs17197087 | 6:31158455:C:T | TCF19 | GTeXv8 Pituitary | 4.58E-22 | T | C | 1.108 | 0.976 | T |
|  | rs143334143 | 6:31153649:G:A | A | rs17197087 | 6:31158455:C:T | TCF19 | Chen CD4Tcells | 1.35E-18 | T | C | 1.82 | 0.976 | T |
|  | rs143334143 | 6:31153649:G:A | A | rs17197087 | 6:31158455:C:T | TCF19 | GTeXv8 Brain Cerebellar Hemisphere | 1.95E-17 | T | C | 1.215 | 0.976 | T |
|  | rs143334143 | 6:31153649:G:A | A | rs17197087 | 6:31158455:C:T | TCF19 | GTeXv8 Brain Hypothalamus | 8.32E-17 | T | C | 1.383 | 0.976 | T |
|  | rs143334143 | 6:31153649:G:A | A | rs17197087 | 6:31158455:C:T | TCF19 | GTeXv8 Muscle Skeletal | 6.93E-14 | T | C | 0.474 | 0.976 | T |
|  | rs143334143 | 6:31153649:G:A | A | rs17197087 | 6:31158455:C:T | TCF19 | GTeXv8 Brain Caudate basal ganglia | 4.73E-10 | T | C | 1.02 | 0.976 | T |
|  | rs143334143 | 6:31153649:G:A | A | rs17197087 | 6:31158455:C:T | TCF19 | GTeXv8 Brain Frontal Cortex BA9 | 6.53E-10 | T | C | 0.961 | 0.976 | T |
|  | rs143334143 | 6:31153649:G:A | A | rs17197087 | 6:31158455:C:T | TCF19 | GTeXv8 Brain Amygdala | 1.14E-09 | T | C | 1.241 | 0.976 | T |
|  | rs143334143 | 6:31153649:G:A | A | rs17197101 | 6:31158632:G:T | TCF19 | Battle Whole Blood | 3.43E-99 | NA | NA | 0 | 0.975 | T |
|  | rs143334143 | 6:31153649:G:A | A | rs17197101 | 6:31158632:G:T | TCF19 | Pala Leucocytes | 2.95E-49 | G | T | -1.648 | 0.975 | T |
|  | rs143334143 | 6:31153649:G:A | A | rs17197101 | 6:31158632:G:T | TCF19 | Davenport Whole Blood | 1.50E-36 | T | G | 0.438 | 0.975 | T |
|  | rs143334143 | 6:31153649:G:A | A | rs17197101 | 6:31158632:G:T | TCF19 | Walsh Whole Blood | 2.54E-15 | NA | NA | 0 | 0.975 | T |
|  | rs143334143 | 6:31153649:G:A | A | rs2233976 | 6:31112217:C:T | TCF19 | Raj CD4Tcells | 2.02E-27 | NA | NA | 0 | 0.872 | T |
| 21q22.1 | rs2236757 | 21:33252612:A:G | A | rs3153 | 21:33237200:G:A | IFNAR2 | Westra Whole Blood | 1.97E-71 | A | G | 17.871 | 0.997 | A |
|  | rs2236757 | 21:33252612:A:G | A | rs574424283 | 21:33232654:T:TAA | IL10RB | Brown LCLs | 5.24E-11 | NA | NA | 0 | 0.997 | TAA |
|  | rs2236757 | 21:33252612:A:G | A | rs12053666 | 21:33246134:G:A | IL10RB | Kasela CD4Tcells | 4.07E-13 | A | G | 7.253 | 0.995 | A |
|  | rs2236757 | 21:33252612:A:G | A | rs2834163 | 21:33248146:G:A | IFNAR2 | GTeXv8 Muscle Skeletal | 1.01E-15 | A | G | -0.239 | 0.883 | G |
|  | rs2236757 | 21:33252612:A:G | A | rs2252639 | 21:33245424:A:G | IFNAR2 | LloydJones Whole Blood | 4.60E-44 | A | G | 0.411 | 0.88 | A |
|  | rs2236757 | 21:33252612:A:G | A | rs17860115 | 21:33230000:C:A | IFNAR2 | Huang LCLs | 1.85E-24 | C | NA | 0.483 | 0.88 | A |
|  | rs2236757 | 21:33252612:A:G | A | rs17860115 | 21:33230000:C:A | IFNAR2 | Fairfax Monocytes naive | 1.84E-20 | NA | NA | 0 | 0.88 | A |
|  | rs2236757 | 21:33252612:A:G | A | rs17860115 | 21:33230000:C:A | IFNAR2 | Huang PBMCs | 5.92E-20 | C | NA | 0.678 | 0.88 | A |
|  | rs2236757 | 21:33252612:A:G | A | rs17860115 | 21:33230000:C:A | IFNAR2 | Hao Lung | 1.00E-16 | C | A | 11 | 0.88 | A |
|  | rs2236757 | 21:33252612:A:G | A | rs2834158 | 21:33244908:T:C | IFNAR2 | Fairfax Monocytes IFN | 2.22E-13 | NA | NA | 0 | 0.88 | T |
|  | rs2236757 | 21:33252612:A:G | A | rs17860115 | 21:33230000:C:A | IFNAR2 | Momozawa Monocytes | 4.09E-11 | A | C | -0.118 | 0.88 | A |
|  | rs2236757 | 21:33252612:A:G | A | rs12482556 | 21:33230629:T:C | IFNAR2 | Momozawa CD4Tcells | 1.54E-16 | C | T | -0.147 | 0.879 | C |
|  | rs2236757 | 21:33252612:A:G | A | rs2236758 | 21:33253108:A:G | IFNAR2 | Battle Whole Blood splice | 1.92E-147 | NA | NA | 0 | 0.878 | A |
|  | rs2236757 | 21:33252612:A:G | A | rs2236758 | 21:33253108:A:G | IFNAR2 | Fehrmann Whole Blood | 3.80E-19 | A | G | 8.94 | 0.878 | A |
|  | rs2236757 | 21:33252612:A:G | A | rs2250226 | 21:33260011:T:C | IL10RB | Westra Whole Blood | 2.51E-13 | C | T | 7.319 | 0.878 | C |
|  | rs2236757 | 21:33252612:A:G | A | rs17860241 | 21:33258828:A:G | IFNAR2 | Brown LCLs | 1.17E-30 | NA | NA | 0 | 0.876 | G |
|  | rs2236757 | 21:33252612:A:G | A | rs17860241 | 21:33258828:A:G | IFNAR2 | GTeXv8 Adipose Subcutaneous | 3.71E-18 | G | A | -0.287 | 0.876 | G |

|  |  |  |  |  |  |  |  |  |  |  |  |  |
| --- | --- | --- | --- | --- | --- | --- | --- | --- | --- | --- | --- | --- |
| rs2236757 | 21:33252612:A:G | A | rs17860241 | 21:33258828:A:G | IFNAR2 | GTeXv8 Esophagus Mucosa | 1.49E-16 | G | A | -0.292 | 0.876 | G |
| rs2236757 | 21:33252612:A:G | A | rs11911133 | 21:33256870:A:G | IFNAR2 | Vosa Whole Blood | 2.09E-280 | G | A | -35.782 | 0.875 | G |
| rs2236757 | 21:33252612:A:G | A | rs11911133 | 21:33256870:A:G | IFNAR2 | Davenport Whole Blood | 3.37E-16 | G | A | -0.114 | 0.875 | G |
| rs2236757 | 21:33252612:A:G | A | rs9975538 | 21:33254549:C:T | IL10RB | GTeXv8 Muscle Skeletal | 3.91E-13 | T | C | -0.253 | 0.875 | C |
| rs2236757 | 21:33252612:A:G | A | rs6517156 | 21:33261740:C:G | IL10RB | Vosa Whole Blood | 1.16E-117 | G | C | 23.06 | 0.862 | G |
| rs2236757 | 21:33252612:A:G | A | rs6517156 | 21:33261740:C:G | IL10RB | Joehanes Whole Blood | 5.21E-41 | C | G | -0.038 | 0.862 | G |
| rs2236757 | 21:33252612:A:G | A | rs6517156 | 21:33261740:C:G | IFNAR2 | Lappalainen LCLs | 7.65E-36 | NA | NA | 0 | 0.862 | G |
| rs2236757 | 21:33252612:A:G | A | rs6517156 | 21:33261740:C:G | IL10RB | GTeXv8 Cells Cultured fibroblasts | 1.82E-29 | G | C | -0.264 | 0.862 | G |
| rs2236757 | 21:33252612:A:G | A | rs6517156 | 21:33261740:C:G | IFNAR2 | Brown Adipose | 2.87E-25 | NA | NA | 0 | 0.862 | G |
| rs2236757 | 21:33252612:A:G | A | rs6517156 | 21:33261740:C:G | IL10RB | GTeXv8 Brain Cerebellum | 4.75E-25 | G | C | 0.687 | 0.862 | G |
| rs2236757 | 21:33252612:A:G | A | rs6517156 | 21:33261740:C:G | IL10RB | GTeXv8 Brain Cerebellar Hemisphere | 4.35E-17 | G | C | 0.586 | 0.862 | G |
| rs2236757 | 21:33252612:A:G | A | rs6517156 | 21:33261740:C:G | IL10RB | GTeXv8 Brain Cortex | 2.64E-12 | G | C | 0.427 | 0.862 | G |
| rs2236757 | 21:33252612:A:G | A | rs1131668 | 21:33262573:G:A | IFNAR2 | Walsh Whole Blood | 1.60E-30 | NA | NA | 0 | 0.86 | A |
| rs2236757 | 21:33252612:A:G | A | rs1131668 | 21:33262573:G:A | IFNAR2 | Alasoo Macrophages naive | 1.14E-12 | NA | NA | 0 | 0.86 | A |
| rs2236757 | 21:33252612:A:G | A | rs1131668 | 21:33262573:G:A | IFNAR2 | Alasoo Macrophages IFNg | 3.25E-10 | NA | NA | 0 | 0.86 | A |
| rs2236757 | 21:33252612:A:G | A | rs3216172 | 21:33262748:G:GT | IL10RB | GTeXv8 Nerve Tibial | 1.62E-14 | GT | G | 0.344 | 0.857 | GT |

\* LD estimated based on individuals of European ancestry from the 1000 Genomes project

**Supplementary Table 18.**
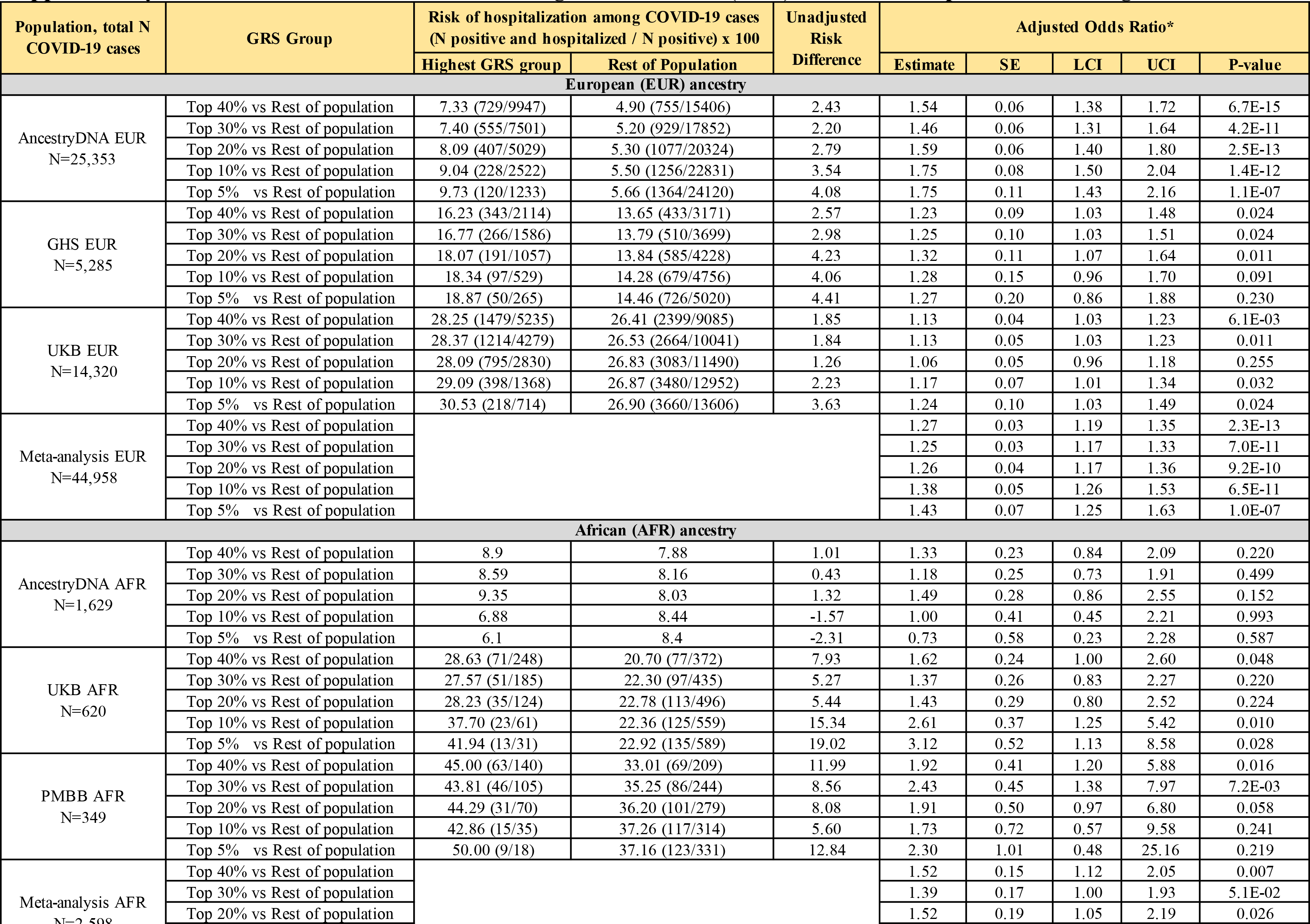
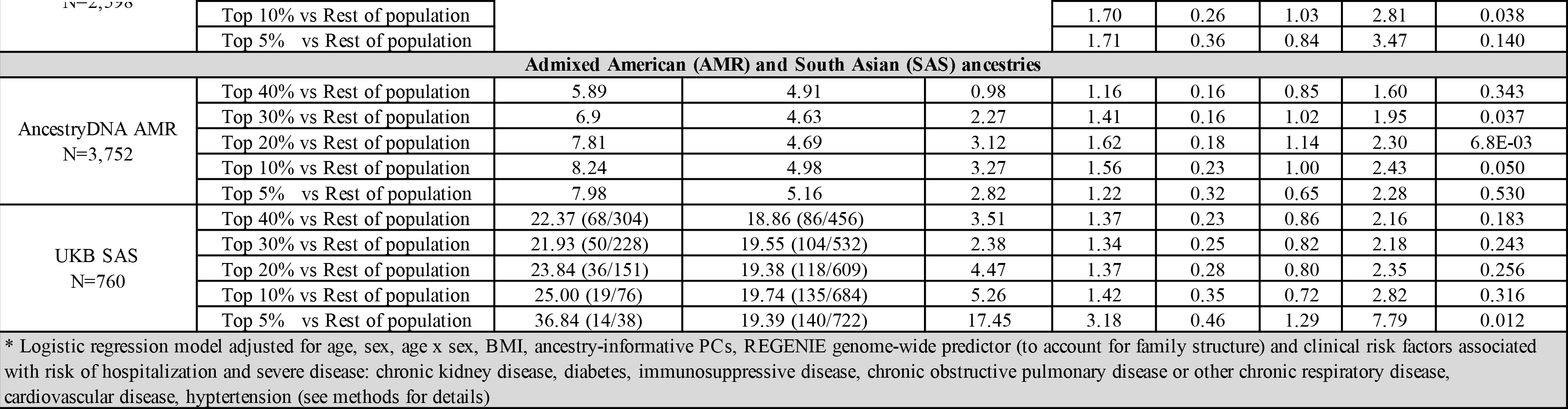
Association between a 6-SNP genetic risk score (GRS) and risk of hospitalization among COVID-19 cases.

**Supplementary Table 19.**
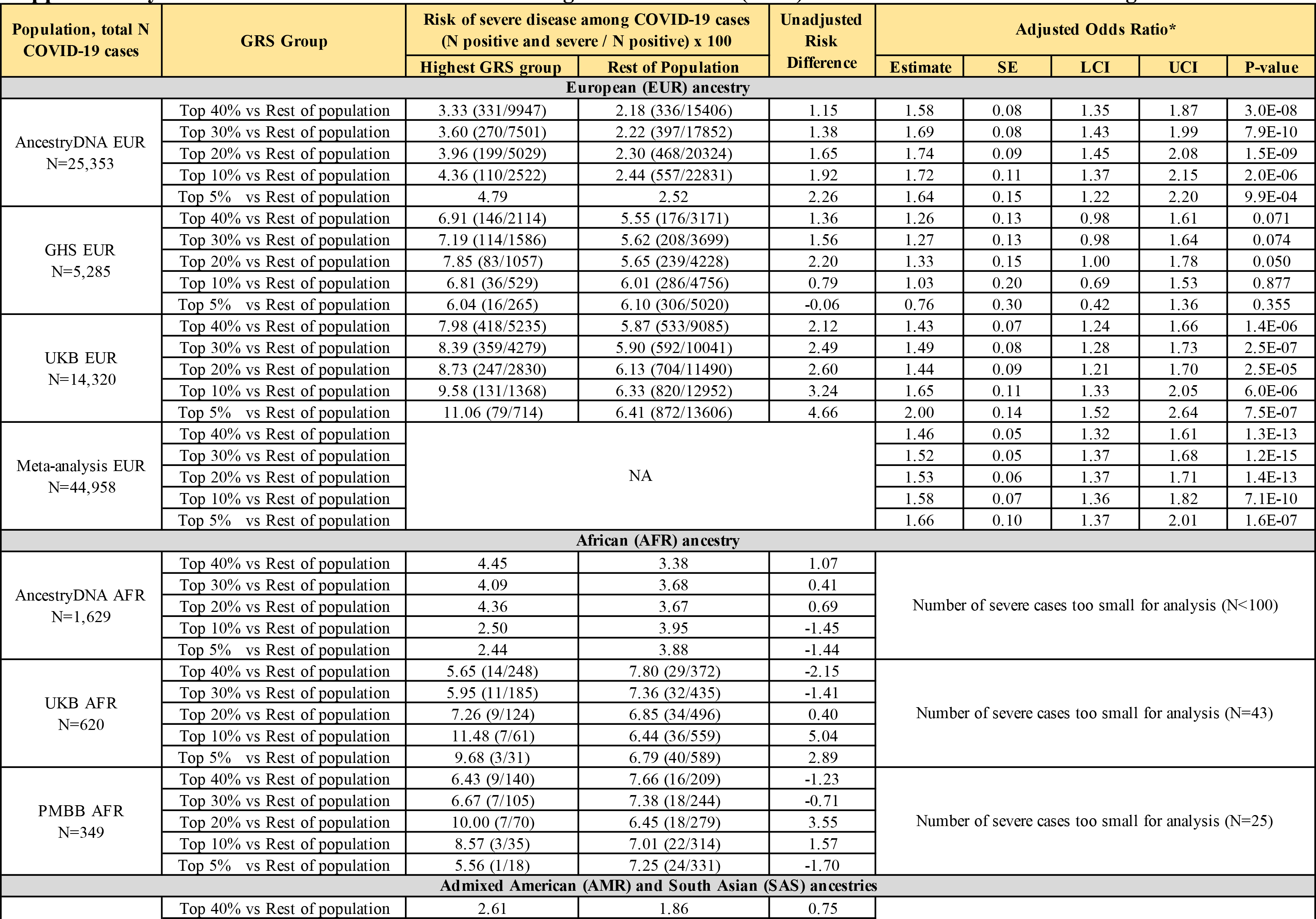
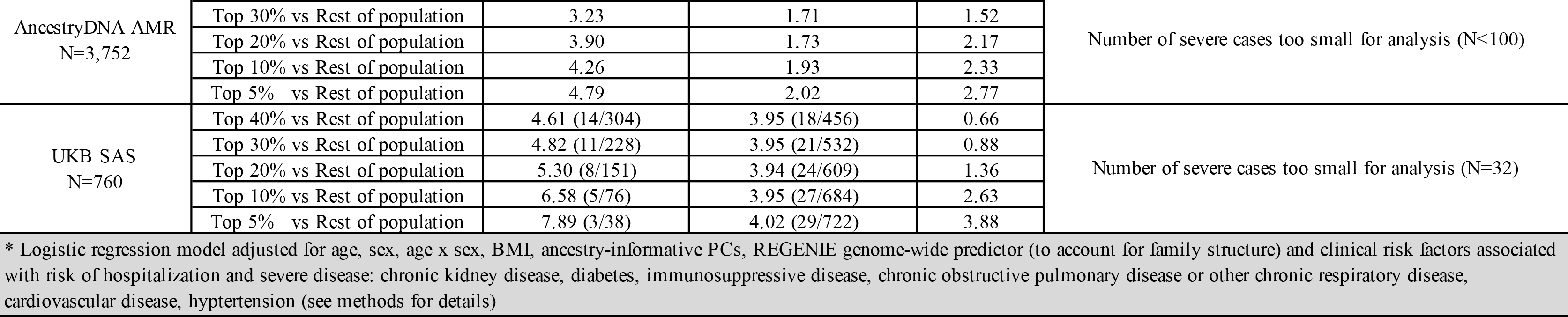
Association between a 6-SNP genetic risk score (GRS) and risk of severe disease among COVID-19 cases.

**Supplementary Table 20.**
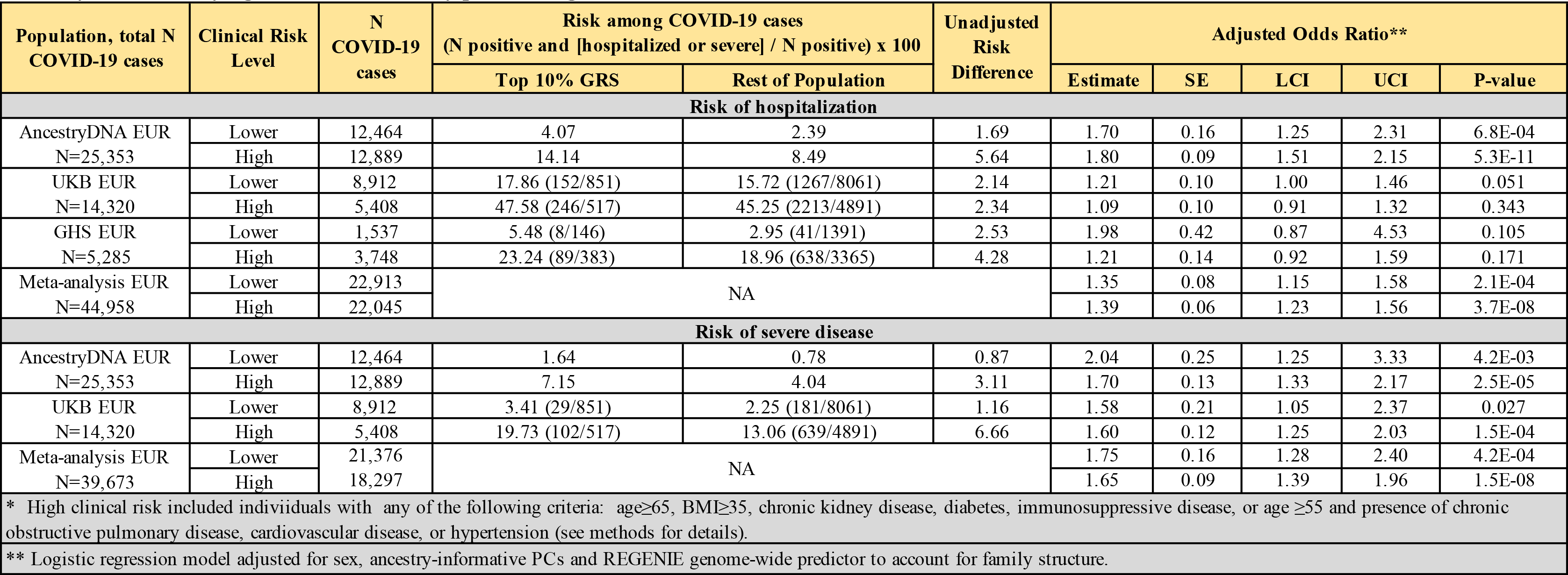
Association between a 6-SNP genetic risk score (GRS) and risk of hospitalization and severe disease in individuals of European ancestry, after stratifying COVID-19 cases by pre-existing clinical risk factor status for severe COVID-19.

